# Synergistic impact of CAG intermediate alleles in the *HTT* gene and microRNA dysregulation exacerbates spliceosome impairment and accelerates Tau pathology in the caudate nucleus of late-onset Alzheimer’s disease

**DOI:** 10.1101/2025.01.19.25320764

**Authors:** Juan Castilla-Silgado, Sergio Perez-Oliveira, Paola Pinto-Hernandez, Manuel Fernandez-Sanjurjo, Maria Daniela Corte-Torres, Eduardo Iglesias-Gutierrez, Manuel Menendez-Gonzalez, Victoria Alvarez, Cristina Tomas-Zapico

**Affiliations:** Department of Functional Biology (Physiology), University of Oviedo, 33006, Oviedo, Spain; Instituto de Investigación Sanitaria del Principado de Asturias (ISPA), 33011, Oviedo, Spain; Department of Neurology, Hospital Universitario Central de Asturias (HUCA), 33011, Oviedo, Spain; Laboratory of Genetics Hospital Universitario Central de Asturias, 33011, Oviedo, Spain; Department of Medicine, University of Oviedo, 33006, Oviedo, Spain; Asociación Parkinson Asturias 33011, Oviedo, Spain; Biobank of Principado de Asturias, Hospital Universitario Central de Asturias (HUCA), 33011, Oviedo, Spain

**Keywords:** CAG repeats, miRNA, proteinopathies, tauopathies, neurodegeneration, splicing factors, neurofibrillary tangles

## Abstract

**Background:** Late-onset Alzheimer’s disease (LOAD) accounts for more than 95% of AD cases. Previously, we have described that 6% of AD patients present CAG intermediate alleles in the huntingtin gene (*HTT IA*s). The caudate nucleus, the most affected region in Huntington’s disease, is highly sensitive to these *HTT* CAG expansions, as they can induce epigenetic changes, including altered microRNA profiles. All this implies a potential source of gene expression deregulation, affecting disease onset and/or progression in LOAD patients with *HTT IAs*.

**Methods:** We genotyped *HTT* CAG repeats and Apolipoprotein E (*APOE*) in *postmortem* brain frozen samples from 323 LOAD patients and 335 healthy controls. From them, we selected caudate samples of *HTT IA* carrier and non-carrier LOAD patients and controls, with neuropathological study, and performed next-generation microRNA sequencing, *in silico* target prediction and pathway analysis, followed by molecular and histopathological studies.

**Results:** Our study revealed that the presence of *HTT IAs* decreases survival in LOAD patients after disease onset. MicroRNA profiles in the caudate nucleus are altered in all LOAD compared to the control cases but are more pronounced in *HTT IAs* carriers. *In silico* analysis suggests that the microRNAs expressed differentially in *HTT IAs* carriers regulate key components of the spliceosome, affecting splicing factors of the SRSF family or the nuclear FUS-SFPQ complex, which was confirmed by different techniques. This leads to an increase in Tau 3R protein, conducting to a higher presence of ghost tangles, the last state of neurofibrillary tangles, in LOAD patients with *HTT IAs*. In addition, they also present a higher number of HTT-positive neurons in a CAG repeat expansion-dependent manner.

**Conclusions:** Our findings demonstrate a synergistic effect of *HTT IAs* and miRNAs deregulation in the evolution of tau pathology, which could be related to an accelerated misprocessing and subsequent aggregation of the Tau 3R isoform, favoring a subsequent faster disease progression. The incorporation of genetic screening for *HTT* alleles into clinical practice would allow a more accurate classification of LOAD patients, facilitating the design of personalized therapeutic interventions and improving the prospects for the management of this debilitating disease.

## Introduction

Many neurodegenerative diseases (NDs) are proteinopathies, characterized by the accumulation of one or more aberrant proteins, with distinctive clinical symptomatology. This is the case of tauopathies, synucleinopathies and polyglutaminopathies. However, there is neuropathological evidence suggesting that proteinopathies are often not pure, but additional proteins may accumulate as copathologies, especially in the more advanced stages of primary ND [1, 2]. Although the specific cause of this phenomenon is not clear, in some cases its origin could be genetic. For example, previous results from our group suggest that the presence of intermediate alleles in the huntingtin gene (*HTT)*, implicated in Huntington’s disease (HD), may play a role in the risk of tauopathies such as Alzheimer’s disease (AD) and frontotemporal dementia (FTD), as well as in synucleinopathies [3–6].

HD is caused by the expansion of the CAG trinucleotide repeat in exon 1 of the *HTT* gene, resulting in an enlarged polyglutamine tract (polyQ) in the HTT protein. When the number of polyQ is greater than 35, an aggregation-prone mutant HTT (mHTT) is produced. HD develops with full penetrance at 40 repeats and above, and the age of disease onset is inversely proportional to the polyQ number. However, between the range of 36-39 repeats is considered incomplete penetrance, whereby HD symptoms develop very late or not at all. Interestingly, *HTT* CAG repeats within the 27-35 range, considered intermediate alleles (*HTT IA*s), have been described to be genetically unstable and prone to expand in offspring, as are pathogenic forms [7, 8]. In fact, the prevalence of *HTT IA*s in the general population can reach 8.7%, rising to 11.6% in relatives with HD [9]. There is substantial evidence supporting interactions between mHTT and Tau protein, one of the histopathological hallmarks of AD, with HD even being proposed as a 4R tauopathy [10]. In addition, a neuropathological study in HD elderly patients showed evidence of copathology with AD in 82% of cases that, at the clinical level, showed prominent dementia [11]. Conversely, there is also evidence suggesting HTT copathology in AD patients, as diffuse HTT accumulation has been found in memory-associated hippocampal regions and in layer III of the frontal cortex above that observed in healthy controls [12].

Therefore, the role of HTT as a modulator of AD neurodegeneration is possible. Indeed, our previous studies indicate that the frequency of *HTT IA*s in AD patients reaches 6%, compared with 2.9% observed in controls [3, 5, 6]. Although the presence of *HTT IA*s need not give rise to mHTT, they can affect at the molecular and cellular levels a multitude of processes important for neuronal physiology. Moreover, the number of CAG repeats in the *HTT* gene has been associated with changes in gene expression in a repeat length-dependent manner, with the striatum being the most sensitive region [13], and such alteration could be given by modifications in epigenetic modulators, such as microRNAs (miRNAs) [14].

In this study, we focused on the effect of *HTT IA*s in patients with late-onset AD (LOAD), as it accounts for more than 95% of AD patients. To determine how these *HTT IA*s might be affecting disease progression, we considered miRNA profiling, as it offers the possibility to accurately identify pathophysiological events affecting neurodegeneration, paving the way towards the development of earlier, more effective and personalized therapeutic interventions. We analyzed a group of healthy subjects and LOAD patients, carriers and non-carriers of *HTT IA*s. Our clinical data suggest that the pathology progresses more rapidly in patients carrying *IA*s. Sequencing of miRNAs in the caudate nucleus allowed us to determine that *HTT IA*s further modifies the LOAD-associated miRNA profile, many of them being involved in the regulation of the nuclear spliceosome machinery, through inhibition of the arginine/serine-rich splicing factor (SRSF) family. Several members of this family, implicated in the splicing of exon 10 (E10) of the microtubule-associated protein Tau (*MAPT*) gene, showed lower mRNA levels in LOAD patients carrying *HTT IA*s. In fact, we confirmed this 4R/3R imbalance by the increased formation of the nuclear complex formed by the sarcoma fused protein (FUS) and proline-glutamine-rich splicing factor (SFPQ), which favors the increase of Tau 3R. Finally, we also found an increased proportion of Tau 3R-rich ghost tangles, a hallmark of late-stage AD neuronal degeneration, in the caudate nucleus of LOAD patients with *HTT IA*s. Our results suggest an important role of *HTT IA*s as risk/progression biomarkers in AD that will also allow us to identify new therapeutic targets, including the *HTT* gene itself, for early neuroprotective strategies. Moreover, it also provides a genetic biomarker in clinical practice, which can be determined with non-invasive tests, quickly and at low cost.

## Materials and methods

### Subjects

This study included brain samples of LOAS patients (n= 323). Material and data supply for patients included in this study were provided by the HCB-IDIBAPS Biobank (B.0000575), integrated in the platform ISCIII Biobanks and Biomodels, and by the Principado de Asturias BioBank (PT23/0077), part of the Spanish National Biobanks and Biomodels Network financed with European funds. Both materials and data were processed following standard operating procedures with the appropriate approval of the Ethics and Scientific Committees. The neuropathological LOAD diagnosis was performed according to standard international consensus criteria [15]. Gender, onset age, age at death, *postmortem* interval (PMI), Braak stages and additional neuropathological findings were collected for each subject. Furthermore, we included a control cohort consisting of 335 healthy controls of whom 40 were brain donors with a neuropathological study. The remaining participants comprised aged controls recruited from Hospital Universitario Central de Asturias (Oviedo, Spain). Gender and age of death were collected. All the participants or legal representatives gave written informed consent to participate in the study. All procedures have been approved by the Ethics Committee of the Principality of Asturias (CEImPA n° 2022.266).

### DNA extraction and genotyping

Genomic DNA was isolated from caudate brain nucleus of LOAD patients and 40 healthy donors and peripheral blood leukocytes (control population) following standard procedures. For the studies of miRNA profile and histopathological studies in the caudate nucleus a group of healthy controls (N= 5), *HTT IA* carrier LOAD (N= 14), and LOAD non-carriers (N= 13) were selected adjusted by sex and age (Supplementary Table 1). DNA extraction from brain and blood samples and genotyping of *HTT* gene CAG repeat number and *APOE* isoforms (SNPs rs7412+ rs429353589) were done as previously described [5].

### Caudate nucleus RNA-sequencing for the analysis of microRNAs

Under cold conditions, 5 mg of frozen tissue was homogenized using Quiazol lysis reagent (QIAGEN, Germany; Cat. No. 79306). RNA extraction, including miRNAs, was subsequently performed using the MIRNeasey micro kit (QIAGEN, Cat. No 74004) following manufacturer’s instructions. The concentration and purity of the extracted RNA was determined on a NanoDrop One 2000c (Thermo Fisher Scientific, USA). The Agilent 2200 TapeStation system was used to analyze the integrity in each sample through the RNA integrity number (RIN) (Supplementary Table 2). Samples were stored at – 80 °C until use.

The RNA-Seq procedure was performed by Seqplexing (Sequencing Multiplex S.L., Spain), using the NovaSeq X sequencing platform (Illumina), paired-end 2×150bp, from 200 ng of total RNA. After generation of miRNA libraries, the quality was evaluated using the QIAxcel Advanced System (QIAGEN). The data obtained from sequencing were generated as raw data in FASTQ format, determining the quality with the FastQC tool. Complementarily, the software miRTrace was used to evaluate the type of RNA found in each sample, allowing to distinguish between miRNAs, rRNA, tRNA and artifacts. From these data, miRNAs were selected and subjected to bioinformatics analysis for the identification of the miRNAs of interest. For this purpose, different criteria were applied to select the miRNAs that became valid: 1) miRNAs in which the loss of data in subjects within the group did not exceed 50%; 2) miRNAs in which their expression was above 800 reads per million (RPM), since they are those that will have a potentially relevant implication on the biological pathways. All raw individual count data for each raw subject are available on ZENODO, with the accession number “11185109” (https://doi.org/10.5281/zenodo.11185109).

### *In silico* prediction of target genes and pathway analysis of miRNAs

*In silico* computational studies were performed to analyze the identified miRNA and determine the targeted pathways by these miRNAs. For each one, experimentally validated targets were retrieved from the miRTarBase and miRWalk databases, with a specificity score greater than 0.8 [16]. The validated gene targets were used in the DIANA TOOLS tool miRPath v.4 [17], and, determined by the Kyoto Encyclopedia of Genes and Genomes (KEEG), the name of the pathways in which those miRNAs are involved, and their degree of significance were extracted. In addition, miRWalk target analysis for Gene Ontology and gene-gene interactions was used. The data downloaded from miRWalk was used for the representation of such interactions with Cytoscape software. Finally, these data were also subjected to simulation with the PantherDB 18.0 tool to obtain the analysis of the classes of protein encoded by these genes, the main molecular function in which they participate, and the cellular compartments in which they show greater representation.

### Gene expression analysis by qPCR

500 ng of total RNA extracted from caudate nucleus were used for cDNA synthesis using the StaRT reverse transcription kit (AnyGenes, France), following the manufacturer’s instructions. The conditions used were: 10 min at 25 °C, 120 min at 37 °C, and 5 min at 85 °C. Quantification was performed from cDNA using the Perfect Master Mix SYBR Green Kit (AnyGenes) on the 7900HT rapid real-time PCR system (Applied Biosystem). The primers for the *MAPT*, *Tau 4R* and *Tau 3R* genes analyzed were specifically designed by Eurogentec (Belgium). *GAPDH* and *β-ACTIN* were used to perform the internal normalization of the results, and the one with the most stable values for our dataset was chosen *a posteriori*, using the RefFinder tool.

Both the specific pairs of primers with pre-designed sequence and those purchased in a validated form the manufacturer (AnyGenes), as well as the amplification conditions for the PCR cycles are listed in Supplementary Tables 3 and 4. The miRNA levels are represented as relative quantification (2^-ΔΔCt^), as described in [18].

### Western blot

Human caudate tissue (30 mg) was homogenized in lysis buffer (20 mM HEPES pH 7.4, 100 mM NaCl, 50 mM NaF, 5 mM EDTA, 1% Triton X-100) with protease inhibitors (cOmplete™, Mini Protease Inhibitor Cocktail; Roche, Switzerland) in a glass homogenizer on ice. Subsequently, the samples were centrifuged for 10 min at 12 000’ g at 4 °C and the supernatant was collected for analysis. Protein concentration was determined with the NanoDrop One 2000c spectrophotometer (Thermo Fisher Scientific).

Fifty micrograms of total protein were loaded onto 4-12% SDS-polyacrylamide gels and transferred to PVDF membranes (0.2 µm, Amersham^TM^ Hybond; Cytiva), which were then blocked in TBS-T (Tris-buffered saline, 1% Tween-20) supplemented with 5% bovine serum albumin (BSA). Membranes were incubated with primary antibodies, (see Supplementary Table 5), overnight at 4 °C, and, after TBS-T washes, with HRP-conjugated goat anti-mouse IgG (1:20000, ab97040, Abcam, UK) or anti-rabbit IgG (1:20000, ab7090, Abcam) for 1h, room temperature (RT). Protein detection was performed with Amersham^TM^ ECL^TM^ Prime Western Blotting Detection Reagent; Cytiva). Anti-GAPDH antibody (1:500, sc-32233, Santa Cruz Biotechnologies, USA) was used as loading control, overnight at 4 °C.

### Immunohistochemistry and immunofluorescence

For expression analysis by immunohistochemistry, the EnVision FLEX+ kit (Agilent-Dako, K8002; Agilent, USA) and the Dako Autostainer system were used. Paraffin-embedded tissues (5 µm) were deparaffinized, rehydrated and unmasked epitopes by heat induction (HIER) at 95 °C for 20 min. In the PT-LINK pretreatment module (Dako), a pH 6 solution (K8005, Agilent-Dak) was used for Huntingtin and Tau 3R detection, whereas a pH 9 solution (K8004; Agilent-Dako) was applied for Tau 4R.

Endogenous peroxidase activity was blocked with EnVision™ FLEX Peroxidase-Blocking Reagent (SN801; Agilent-Dako) for 5 min. Sections were incubated with Tau-3R (1:400, clone 8E6/C11, 05-803, Millipore, USA), Tau-4R (1:100, clone: 1E1A6, 05-804, Millipore), and Huntingtin (1:50, clone: EM-48, MAB5374, Millipore) primary antibodies diluted in EnVision™ FLEX Antibody Diluent (K8006; Agilent-Dako) for 20 min at RT, and overnight at 4 °C, respectively. The signal was detected using diaminobenzidine chromogen as substrate on Dako EnVision™ FLEX /HRP (DM827+SM803; Agilent-Dako). Sections were counterstained with hematoxylin (K8008; Agilent-Dako). Negative controls were processed omitting the primary antibody. Finally, sections were dehydrated and mounted with permanent medium (Agilent-Dako mounting medium, CS703).

Images were acquired by means of the NanoZoomer-SQ Digital slide scanner (C13140-01, Hamamatsu, Germany) and then quantified using QuPath-0.4.2 software. Positive signal in caudate nucleus neurons was counted in 5 different areas of the slides and then averaged for each mm^2^.

For double immunofluorescence, after antigen retrieval, the sections were washed with distilled water (H2Od). They were then rinsed three times with 0.1M monophosphate buffer and once with PBS for 10 min. Subsequently, the sections were incubated with blocking solution for 45 min (containing 1% BSA and 1% Triton X-100 in PBS). The primary antibody was incubated overnight at 4 °C in blocking solution (supplemented with normal serum); anti-SFPQ (1:100, WH0006421M2; Sigma-Aldrich, USA), and anti-FUS (1:100, A300-293A; Bethyl laboratories, USA). The next day, the sections were washed with PBS and then with H2Od. The secondary antibodies, donkey anti-mouse Alexa-488 and donkey anti-rabbit Alexa 594 (1:1000, Invitrogen; Thermo Fisher Scientific), were incubated 2 h at RT. Finally, the sections were treated with Sudan Black for 10 min and the nuclei were stained with DAPI included in the mounting medium (Fluoroshield^TM^, Sigma-Aldrich).

Images were acquired on a Leica TSC-SP8X spectral confocal microscope (Leica DMI8 microscope) with excitation lines between 470-670 nm and PLA APO 20X/0.75 IMM CORR CS2 or PLA APO 40X /1.30 CS2 oil-immersion objective (Leica Microsystems, Germany). Images were acquired with Leica Application Suite X software (version 1.8.1, Copyright 1997-2015; Leica Microsystems) and processed with LAS_X_SMALL (version 1.0.0) and Uniovi Fiji Confocal/ImageJ (version 1.6) software.

### Nuclear colocalization analysis

To perform a quantitative analysis of the colocalization status between FUS and SFPQ in neuronal nuclei, all sections were analyzed under the same configuration and laser intensity in the confocal microscope, acquiring random images in two regions of the caudate nucleus and six Z-planes, with 1 µm jumps, traversing the entire volume of the nucleus at a magnification of 630’. Cells of interest were identified by nucleus size (∼ 10-12 µm in diameter) and nuclei with a diameter < 8 µm were discarded from the measurements. Measurements were performed on 30 randomly selected nuclei in each section. To study colocalization, all images were analyzed separately using the *Coloc 2* plug-in in FIJI software. Threshold Mander’*s* coefficients (tM1 and tM2) and Pearson’s correlation coefficient (R) were calculated to measure the strength of colocalization and correlations between fluorescence channels.

### Proximity ligation assay (PLA)

The proximity ligation assay for the detection of protein complexes was carried out as described [19]. Briefly, paraffin-embedded brain tissue was rehydrated and subjected to antigenic unmasking in pH 6.0 solution at 95 °C for 20 min. Subsequently, plasma membranes were permeabilized by incubation with 0.1% Triton X-100 in PBS for 15 min. The sections were blocked for 45 min with a specific blocking solution (DUO92009, Sigma-Aldrich). For detection of protein complexes, samples were incubated with anti-FUS (1:300, A300-293A, Bethyl laboratories) and anti-SFPQ (1:100, WH0006421M2, Sigma-Aldrich) primary antibodies in dilution buffer at 4 °C overnight. After antibody recovery, the sections were washed twice with 1× buffer A (DUO82049, Sigma-Aldrich) and incubated for 1 h at 37 °C with PLUS (DUO92002, Sigma-Aldrich) and MINUS (DUO92004, Sigma-Aldrich) probes diluted 1:5. The ligation procedure was then performed by incubation with ligase diluted 1:40 in ligation buffer (DUO92014, Sigma-Aldrich) at 37 °C for 30 min. Probes were amplified by adding polymerase diluted 1:80 in amplification buffer, incubating at 37 °C for 100 min. Finally, sections were washed with 1× and 0.01× buffer B (DUO82049, Sigma-Aldrich), mounted with DAPI mounting medium (DUO82040, Sigma-Aldrich) and stored until microscopic observation.

To determine the signal intensity, all sections were analyzed under the same configuration and laser intensity on the confocal microscope, acquiring random images in 5 Z-planes, with 1 µm jumps, scanning the entire volume of the nucleus at a magnification of 630x. Cells of interest were identified by nucleus size (∼ 10-12 µm in diameter) and nuclei with a diameter < 8 µm were discarded from the measurements. These measurements were performed on 17 randomly selected nuclei in each section. To measure the fluorescence intensity in each nucleus, the maximum projection was made in each image, obtaining the joint intensity in each Z layer. Next, the *stack profile* tool in LAS X *software* (Leica Application Suite X, Leica Microsystems) was used and the nuclei were individually circled to obtain the mean fluorescence intensity (in gray values between 0-255). Finally, the fluorescence measurement was referenced with respect to the area of each nucleus and the data were averaged to obtain the total intensity in each subject.

### Statistical analysis

Graphs were produced using GraphPad Prism 10.2.0 (GraphPad, USA) and SRPlot module [20] and statistical analyses were performed using IBM SPSS Statistic 27.0 software (IBM, USA). Categorical variables were described as frequencies and percentages. For between-group comparisons of the frequencies of *HTT IAs*, *APOE* alleles or gender frequency, Chi-square and Fisher tests were performed. To determine the relationship between the presence of *HTT IAs* and the probability of survival in the LOAD groups, the Kaplan-Meier estimation method and the log-rank test were used. Survival rate was calculated as the time elapsed from pathology diagnosis to age at death (event = 1 per subject). In addition, multivariate Cox-PH regression models were run to identify predictors of survival risk, such as age or the presence of *HTT IAs*. Mean and standard deviation or median and interquartile range, as appropriate, were used to describe quantitative variables. Kolmogorov-Smirnov and Shapiro-Wilks normality tests (for *N* < 50) were performed and the corresponding parametric or nonparametric tests were then applied. To compare age at onset, age at death, disease duration, and miRNAs expression levels between LOAD groups, Student’s t-test or Mann-Whitney U-test was performed, as appropriate. Also, the parametric analysis of variance (ANOVA) test, followed by Dunnett’s comparison, or the nonparametric Kruskal-Wallis’ test, followed by Dunn’s test, was applied when comparisons were made with the control group. To determine whether miRNAs levels correlate with the recording of different clinical variables, a Spearman correlation test was applied. Graphical representation of miRNAs levels in heatmap was performed by normalization through Z-score. For immunohistochemistry and immunofluorescence studies, Student’s t-test or ANOVA, followed by the corresponding *post hoc* test, were applied as appropriate. The threshold for statistical significance was set at *p* < 0.05. The exact levels of the *p-values* are indicated in each figure or additional table, as applicable.

## Results

### The presence of *HTT IAs* modifies LOAD progression

We have previously analyzed the possible influence of the *HTT IAs* on the development of tauopathies, including AD [5]. The results suggested that LOAD patients had a higher frequency of *HTT IAs* than patients with early-onset AD, which could be affecting disease progression. Thus, to further explore the effect of *HTT IAs* on LOAD, we considered for this study only LOAD patients from our previous cohort and a group of healthy subjects [5]. The age of LOAD onset was 75.22 ± 6.02 years and disease duration until death was 9.35 ± 4.86 years, with a higher percentage of women in the LOAD group versus the control (Table 1). The frequency of distribution of the different isoforms for the *APOE* gene was accordance with what was previously described (Table S6; [5]). The presence of subjects carrying *HTT IAs* was higher in the case of LOAD patients versus healthy controls, with a frequency similar to that observed in previous studies (7.12% *vs.* 4.16%; *p-value* = 0.102; Fisher’s test, Table 1) [3, 5]. Moreover, the number of CAG repeats was higher in the long *HTT* allele of LOAD subjects with respect to healthy controls (20.42 ± 3.33 *vs.* 19.61 ± 3.22, respectively; *p-value* < 0.001; Student’s t-test; Table 1 and Figure S1B). Importantly, within the range of IAs in the LOAD group, the most frequent expansion was 27 CAGs, with subjects presenting even 35 CAGs (Figure S2).

**Table 1.**
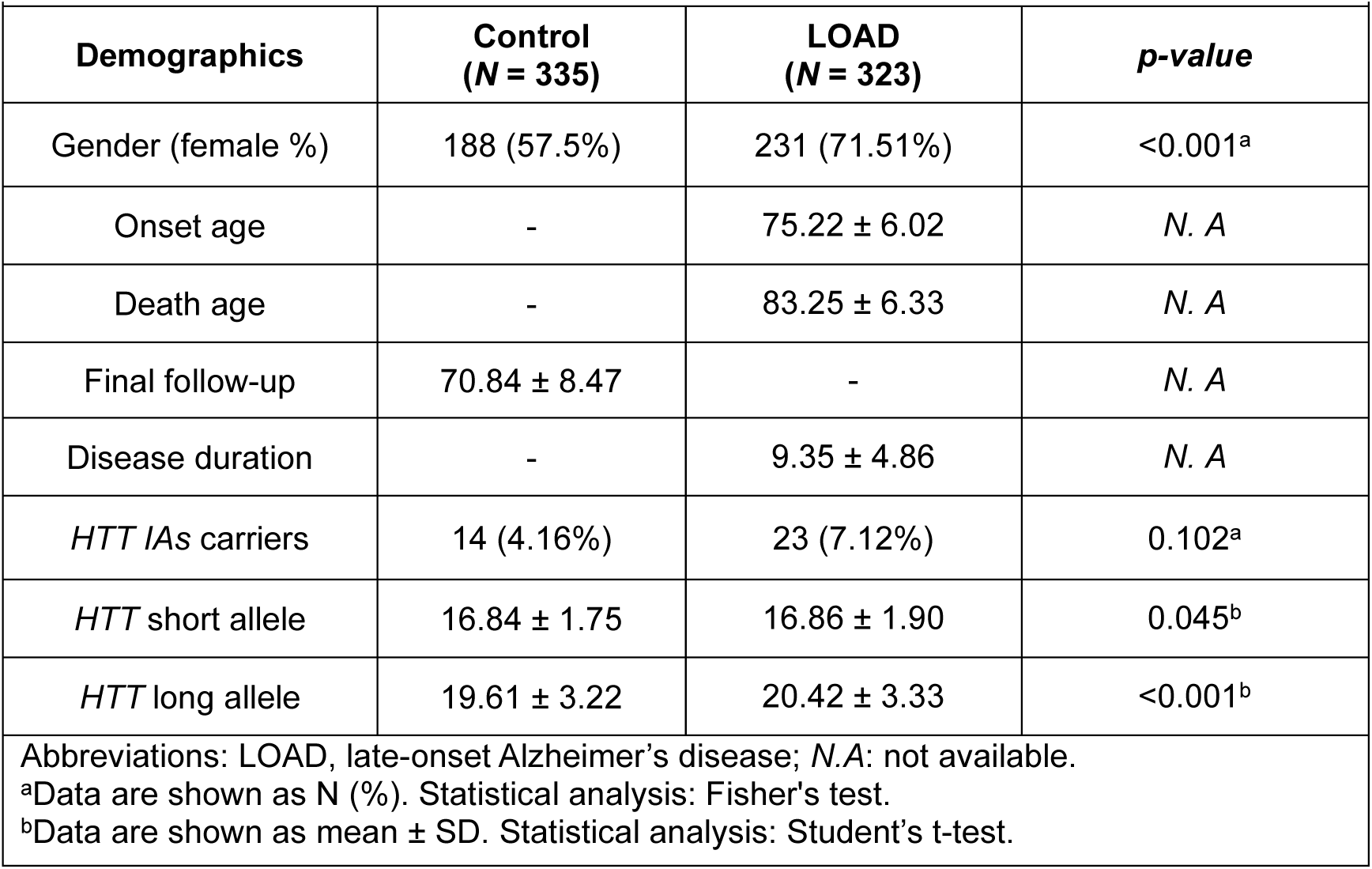
Demographic data of cohorts studied.

To study the effect of the presence of *HTT IAs* specifically in LOAD donors, we divided this group into non-carrier (*N =* 300) and carrier (*N =* 23) patients (Table 2). The number of CAG repeats in the long *HTT* allele was higher in carriers versus non-carriers (29.00 ± 2.37 *vs.* 19.88 ± 2.54 CAG repeats; *p-value* < 0.001, Student’s t-test; Table 2). No significant differences were found in the percentage of females, age at onset, and age at death between the two groups. However, the duration of the disease, after diagnosis, was shorter in carrier subjects with respect to non-carrier (7.40 ± 2.89 years *vs.* 9.50 ± 4.96 years, respectively; *p-value* = 0.053, Student’s t-test; Table 2).

**Table 2.** Demographics data of LOAD cohort studied.

| Table 2. Demographics data of LOAD cohort studied. |  |  |  |
| --- | --- | --- | --- |
| Demographics | *LOAD non-HTT IAs*<br>(*N* = 300) | *LOAD HTT IAs*<br>(*N* = 23) | *p-value* |
| Gender (female %) | 216 (66.87%) | 15 (65.21%) | 0.488<sup>a</sup> |
| Onset age | 75.24 ± 5.98 | 75.75 ± 6.64 | 0.855<sup>b</sup> |
| Death age | 84.33 ± 6.31 | 83.43 ± 6.79 | 0.492<sup>b</sup> |
| Disease duration | 9.50 ± 4.96 | 7.40 ± 2.89 | 0.053<sup>b</sup> |
| *HTT* short allele | 16.82 ± 1.90 | 17.48 ± 1.93 | 0.057<sup>b</sup> |
| *HTT* long allele | 19.88 ± 2.54 | 29.00 ± 2.37 | <0.001<sup>b</sup> |
| <sup>a</sup>Data are shown as N (%). Statistical analysis: Fisher's test. |  |  |  |
| <sup>b</sup>Data are shown as mean ± SD. Statistical analysis: Student's t-test. |  |  |  |

In fact, the survival rate after diagnosis showed a clear reduction of this rate in LOAD donors with *HTT IA*s (*p-value* = 0.012; Kaplan-Meier estimator; Figure 1). To verify this result, logistic regression models were used. The model proposed revealed a significant fit (Cox-Snell R^2^ = 0.01) and a significant association between disease duration and the presence of *HTT IAs* (*p* = 0.012). These results suggest that the *HTT IAs* modifies LOAD progression, decreasing patient survival after disease onset.

**Figure 1.**
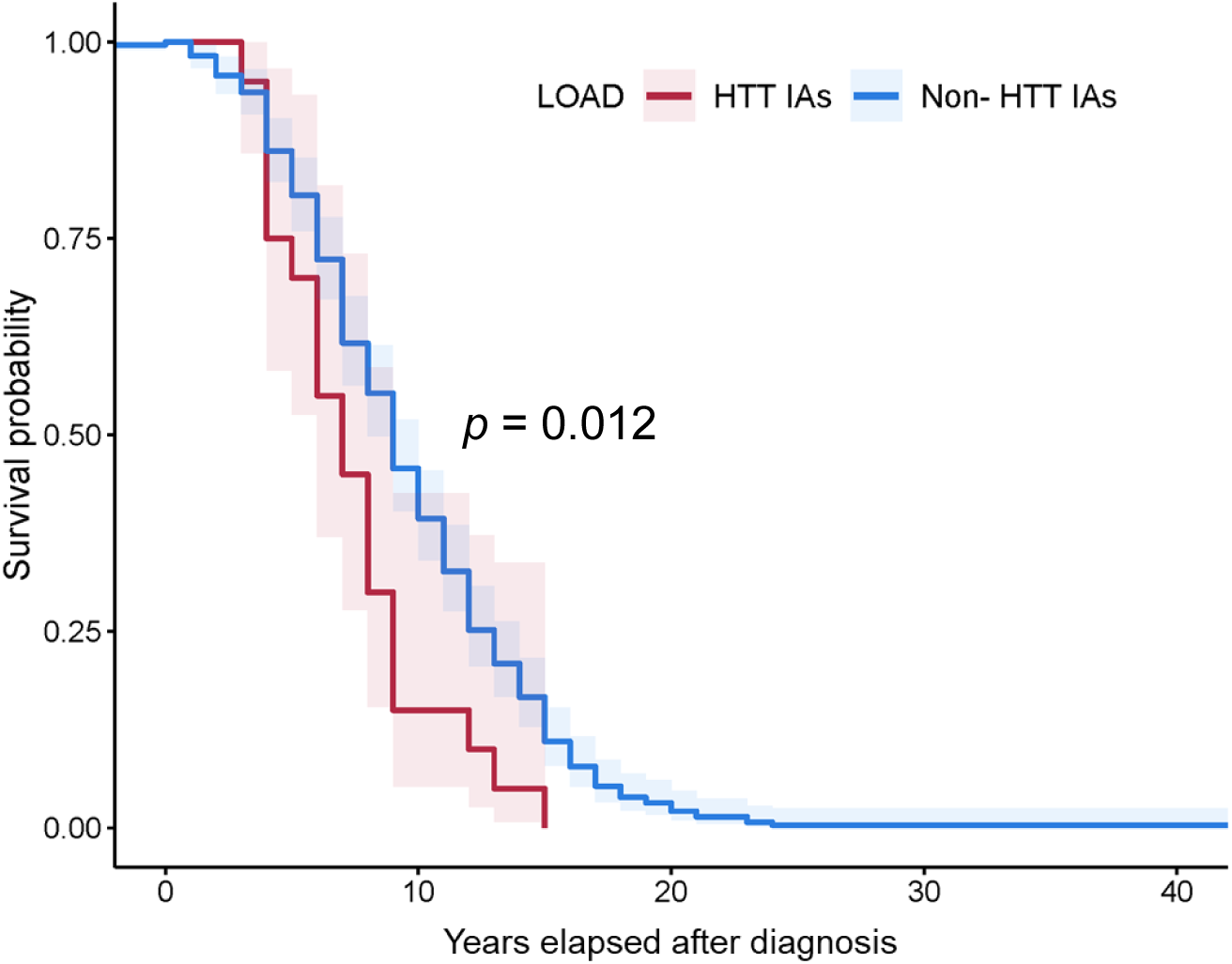
The presence of *HTT IA*s modifies LOAD progression. Comparative analysis of survival function by Kaplan-Meier estimation shows a reduction in survival, after clinical diagnosis of the disease, in the group of LOAD donors with *HTT IAs* compared to non-*HTT IAs*. Statistical significance was calculated by Kaplan-Meier estimator and Log-rank test. *P-value* is indicated in the graph.

### Caudate nucleus miRNA profiling LOAD patients shows size-dependent alterations of the CAG expansion in the *HTT* gene

In murine models of HD, altered expression of the miRNA profile as a function of the number of CAG expansions in the *HTT* gene was observed in different brain regions, being the striatum the most vulnerable area [14]. Therefore, it is possible that the caudate nucleus of LOAD patients carrying *HTT IAs* shows an altered miRNA profile in this region. We performed RNA-Seq on caudate nucleus *postmortem* samples and, after selection (see Materials and Methods section), a total of 33 miRNAs met the requirements. From these, we performed a principal component analysis (PCA), where PC1 and PC2 components explained 57.7 % of the variance observed in the subjects (Table S7). Thirty of the miRNAs were contributing to PC1 (loading coefficient > 0.5), varying together in this component, among which miR-9-5p and miR-30a-5p presented the highest loading coefficient (Table S7). In the case of PC2, only four miRNAs were most correlated with this component, of which three showed a negative correlation, miR-26b-5p, miR-26a-5p and let-7i-5p. Figure 2A shows the graphical representation of PC2 versus PC1. The representatives of the control group displayed low values for the variables that contribute most to these two components, while the patients of the two LOAD groups analyzed showed higher values in both components, being even higher in the LOAD patients carrying *HTT IAs* group, but not enough to make a distinction between the three groups.

**Figure 2.**
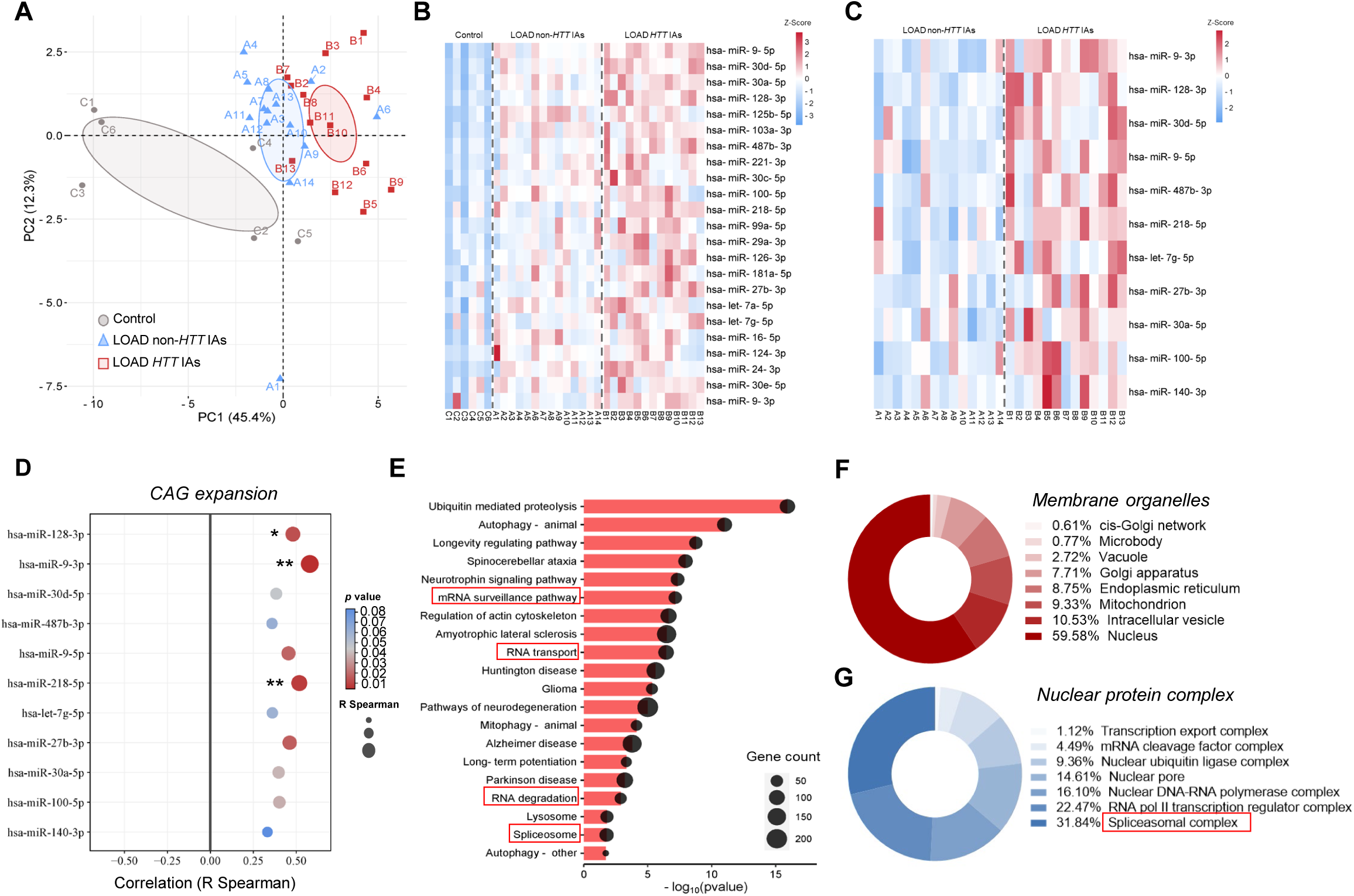
miRNA profile in the caudate nucleus links CAG expansion in *HTT* gene to spliceosome regulation. **(A)** Graphical plot of principal component analysis (PCA) of differentially expressed miRNAs considering the three studied groups. **(B-C)** Heat maps showing miRNAs expression after RNA-seq analysis in caudate nucleus samples from control subjects *vs*. LOAD subgroups **(B)** and considering only LOAD donors (*HTT IAs vs. non-HTT IA* carriers; **C**). Colors scale illustrates the most highly expressed miRNAs from red to blue (*p-value* after Dunnet’s test *vs.* control comparison <0.05; Z-score). **(D)** Spearman correlation between CAG expansion size in *HTT* gene and miRNAs expression. Dot size is proportional to the correlation value and the *p-value* is represented by the color scale in the legend (* *p-value* < 0.05, ** *p-value* < 0.01). **(E)** Diagram KEEG enrichment analysis pathways in which differentially expressed miRNAs target between LOAD subgroups. The size of the circles refers to the number of target genes for the miRNAs within each pathway. **(F-G)** Gene ontology analysis in 13 198 genes identified using Panther DB platform and distribution according to their compartment of action in membrane organelle **(F)** and cellular protein complex in the nucleus **(G)**. Next to each distribution, the percentage frequency of the genes or the number counted, as appropriate, is indicated. The number of genes assigned may be greater than the total recognized, since the same gene may be included in more than one category.

Analysis to determine which of these miRNAs were differentially expressed among the three groups showed that 23 of the 33 miRNAs initially detected were altered in LOAD patients (Table 3), with higher expression levels with respect to the control group (Figure 2B). From the PCA analysis and differential expression results, we observed that the differences regarding control group were higher in LOAD donors with *HTT IAs*. To determine whether this also implies a change in the expression profile with respect to non-carrier patients, we performed a differential analysis of the expression of the 33 miRNAs found considering only these two groups. This revealed that ten of these miRNAs are more expressed in the *HTT IAs* carrier than in the non-*HTT IAs* LOAD patients and, in addition, a new miRNA, miR-140-3p, whose differential expression is specific between the LOAD groups, was identified (Table 4 and Figure 2C). All this suggests that the miRNAs profile would be affected by the pathology itself and that the size of the CAG expansion in the *HTT* gene exacerbates it even more.

**Table 3.** Differentially expressed miRNAs in healthy controls *vs.* LOAD groups.

| miRNAs | Control | LOAD *non-HTT* IAs<br>[*p*-value vs. Control] | LOAD *HTT* IAs<br>[*p*-value vs. Control] |
| --- | --- | --- | --- |
| hsa-miR-9-5p | 16386.6 ± 7417.38 | 32643.36 ± 5013.56<br>[<0.025]* | 39014.65 ± 5207.62<br>[<0.001]*** |
| hsa-miR-30d-5p | 2819.36 ± 1657.27 | 5793.21 ± 1134.76<br>[<0.001]*** | 7545.85 ± 1484.60<br>[<0.001]*** |
| hsa-miR-30a-5p | 3941.43 ± 2223.77 | 7719.60 ± 1488.43<br>[<0.001]*** | 9224.76 ± 1615.98<br>[<0.001]*** |
| hsa-miR-128-3p | 4182.43 ± 2453.05 | 7897.71 ± 1813.42<br>[<0.001]*** | 11928.67 ± 3091.31<br>[<0.001]*** |
| hsa-miR-125b-5p | 1305.32 ± 611.40 | 2751.71 ± 557.81<br>[<0.001]*** | 2583.71 ± 471.96<br>[<0.001]*** |
| hsa-miR-103a-3p | 3531.15 ± 2064.04 | 7974.13 ± 2293.12<br>[<0.001]*** | 9017.74 ± 2049.48<br>[<0.001]*** |
| hsa-miR-487b-3p | 889.57 ± 571.90 | 1477.67 ± 402.52<br>[0.057] | 2171.85 ± 648.72<br>[<0.001]*** |
| hsa-miR-221-3p | 946.90 ± 466.80 | 1827.74 ± 641.08<br>[0.014]* | 2250.74 ± 693.33<br>[<0.001]*** |
| hsa-miR-30c-5p | 5551.12 ± 2778.08 | 9428.42 ± 1717.47<br>[0.005]** | 10713.78 ± 3368.41<br>[0.001]** |
| hsa-miR-100-5p | 5212.68 ± 2435.92 | 9829.22 ± 2232.98<br>[0.095] | 10515.86 ± 3491.47<br>[0.001]** |
| hsa-miR-218-5p | 1495.30 ± 904.06 | 2005.07 ± 656.07<br>[0.173] | 2685.15 ± 433.87<br>[0.001]** |
| hsa-miR-99a-5p | 3093.67 ± 1188.20 | 5094.84 ± 1597.55<br>[0.038]* | 6288.39 ± 1970.75<br>[0.001]** |
| hsa-miR-29a-3p | 1946.82 ± 799.29 | 2779.13 ± 803.40<br>[0.004]** | 3582.45 ± 951.83<br>[0.001]** |
| hsa-miR-126-3p | 3125.08 ± 1920.87 | 5420.40 ± 1409.52<br>[0.065] | 7066.36 ± 2796.03<br>[0.001]** |
| hsa-miR-181a-5p | 1127.99 ± 647.37 | 1832.21 ± 638.10<br>[0.061] | 2354.87 ± 704.51<br>[0.001]** |
| hsa-miR-27b-3p | 2717.82 ± 1559.54 | 3526.19 ± 808.69<br>[0.247] | 4626.31 ± 1250.03<br>[0.004]** |
| hsa-let-7a-5p | 1855.95 ± 964.69 | 3017.19 ± 536.48<br>[0.02]* | 3246.25 ± 798.72<br>[0.005]** |
| hsa-let-7g-5p | 7628.01 ± 4545.65 | 9584.85 ± 1628.66<br>[0.219] | 11765.65 ± 2436.24<br>[0.006]** |
| hsa-miR-16-5p | 1099.82 ± 620.93 | 1895.01 ± 431.61<br>[0.004]* | 1840.18 ± 481.52<br>[0.008]** |
| hsa-miR-124-3p | 3016.37 ± 1393.79 | 6340.28 ± 3470.01<br>[0.007]* | 6776.95 ± 2815.46<br>[0.009]** |
| hsa-miR-24-3p | 4169.52 ± 2147.22 | 11502.07 ± 3279.17<br>[0.001]** | 13890.95 ± 4082.34<br>[0.009]** |
| hsa-miR-30e-5p | 2271.66 ± 1610.20 | 3254.16 ± 869.36<br>[0.129] | 3809.25 ± 1087.95<br>[0.015]* |
| hsa-miR-9-3p | 8417.09 ± 6358.31 | 9191.86 ± 1803.44<br>[0.804] | 12182.08 ± 1913.93<br>[0.034]* |

Data are shown as mean ± SD. Statistical analysis: One-way ANOVA followed by Dunnet's test. Ordered from highest to lowest significance according to LOAD *HTT* IAs. Statistically significance differences, \* *p*<0.05, \*\* *p*<0.01, \*\*\**p*<0.001.

**Table 4.** Differentially expressed miRNAs between LOAD non-*HTT IAs* and LOAD *HTT IAs* patients.

|  |  |  |  |
| --- | --- | --- | --- |
| miRNAs | LOAD non-*HTT* IAs | LOAD *HTT* IAs | *p*-value |
| hsa-miR-9-3p | 9191.86 ± 1803.44 | 12182.08 ± 1913.93 | <0.001*** |
| hsa-miR-128-3p | 7897.71 ± 1813.42 | 11928.67 ± 3091.31 | <0.001*** |
| hsa-miR-30d-5p | 5793.21 ± 1134.76 | 7545.85 ± 1484.60 | 0.002** |
| hsa-miR-9-5p | 32643.36 ± 5013.56 | 39014.65 ± 5207.62 | 0.003** |
| hsa-miR-487b-3p | 1477.67 ± 402.52 | 2171.85 ± 648.72 | 0.003** |
| hsa-miR-218-5p | 2005.07 ± 656.07 | 2685.15 ± 433.87 | 0.004** |
| hsa-let-7g-5p | 9584.85 ± 1628.66 | 11765.65 ± 2436.24 | 0.010* |
| hsa-miR-27b-3p | 3526.19 ± 808.69 | 4626.31 ± 1250.03 | 0.011* |
| hsa-miR-30a-5p | 7719.60 ± 1488.43 | 9224.76 ± 1615.98 | 0.019* |
| hsa-miR-100-5p | 2779.13 ± 803.40 | 3582.45 ± 951.83 | 0.025* |
| hsa-miR-140-3p | 1282.55 ± 228.36 | 1586.29 ± 492.22 | 0.048* |
| Data are shown as mean ± SD. Statistical analysis: Student's *t* -test. Ordered from highest to lowest significance. Statistically significance differences, * *p* <0.05, ** *p* <0.1, *** *p* <0.001 |  |  |  |

Therefore, to explore the role that altered miRNAs may play with this genetic condition in the context of LOAD, we examined the relationship between the 11 differentially expressed miRNAs among both LOAD groups and some recorded clinical characteristics. Age at onset, age at death, duration of disease and Braak stage showed no correlation with the expression of any of the miRNAs (Figure S3). However, when analyzing the influence of the size of CAG expansions (Figure 2D), we observed a positive correlation in only 3 of the 11 miRNAs considered, miR-128-3p (R= 0.57, *p-value*= 0.001), miR-9-3p (R= 0.61, *p-value=* 0.0006), and miR-218-3p (R= 0.51, *p-value=* 0.006). Therefore, our results indicate that the observed alteration in the caudate nucleus miRNAs profile is dependent on the size of the CAG expansions in the *HTT* gene, which may influence the faster progression of the pathology in LOAD donors carrying *HTT IAs*.

### The altered miRNAs by the presence of *HTT IAS* may exert regulation on the spliceosome pathway

An initial approach to the results of our *in silico* study revealed, through Cytoscape plots, that several of the altered miRNAs among the LOAD groups are co-targeted to *MAPT* and *HTT* genes (Figure S4A). Thus, we decided to perform an in-depth exploration analysis of these 11 altered miRNAs, using DIANA-miRPathv4.0 integrated with TargetScanv8.0 and KEEG pathways. A linkage of these miRNAs to a total of 127 pathways were found, of which 20 were related to the brain (Figure 2E), like those found if altered miRNAs versus the control group were considered (Figure S4B). These include pathways related to protein homeostasis mechanisms (ubiquitin-mediated proteolysis and autophagy), NDs (AD, HD, spinocerebellar ataxia, and Parkinson’s disease), and pathways associated with RNA processing, such as the mRNA survival pathway, RNA transport, RNA degradation and spliceosome pathway.

To further investigate these results, we analyzed the genes regulated by the 11 selected miRNAs by gene ontology enrichment analysis using the PantherDB v18.0 database. We identified a total of 13,198 target genes, with 90,057 interactions in different genomic regions. These genes were grouped according to protein class, molecular function and cellular components. The majority corresponded to transcriptional regulators (29 %), protein-modifying enzymes (28 %), genes involved in RNA metabolism (14 %) and modulators of protein binding activity (13 %) (Figure S4C). In terms of molecular function, interactions with cyclic and heterocyclic organic compounds (29 % in both cases) and protein binding (25 %) predominated (Figure S4D). In terms of cellular localization, genes were mainly associated with intracellular structures, with membrane-bound organelles standing out, with 59.58 % linked to the nucleus (Figure 2F). Within the nucleus, most of the genes of interest were associated with nuclear protein complexes (Figure 2G), among which 31.8% corresponded to components of the spliceosome machinery. This complex is essential in the mRNA alternative splicing process, crucial for the proper processing of proteins in the brain. Taken together, the *in silico* analysis suggests a significant relationship between the miRNAs profile found in caudate nucleus and the regulation of the spliceosome.

### miRNAs profiling affects the expression of SRSF splicing factor members

Alterations in the spliceosome pathway have been well documented in AD and other neurodegenerative pathologies, including HD [7, 8], with important effects on *MAPT* gene splicing, being miRNAs most likely involved (reviewed in [21]). The regulation underlying this pathway is extremely complex and, within the factors involved in Tau splicing, the SRSF family stands out [22]. The target’s miRNAs analysis indicated that many of them were, precisely, members of this family, suggesting that the alterations observed in the miRNAs profile may affect *MAPT* gene splicing. Figure 3A shows the complex interaction network between many of the miRNAs analyzed and the mRNA of different SRSF family members (SRSF1, SRSF2, SRSF3, SRSF4, SRSF6, SRSF7, SRSF9, SRSF11) or the *MAPT* gene. The connection between targets suggested a complex regulation by these miRNAs, so the altered profile we have found in LOAD patients can have significant effects on the expression of more than one member of the SRSF family and, by extension, on the levels of Tau isoforms.

**Figure 3:**
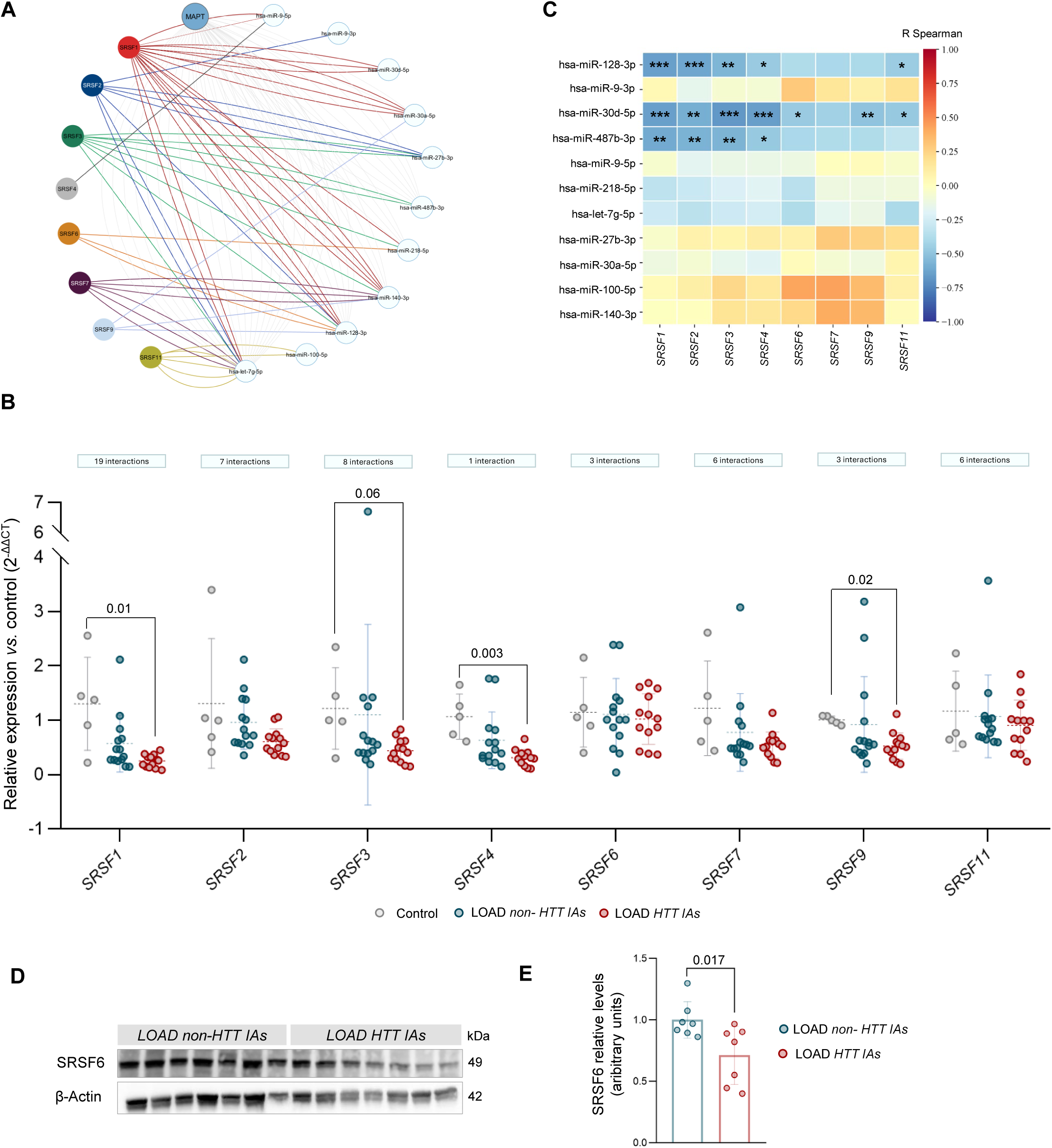
Altered miRNA profile in the caudate nucleus affects the expression of SRSF splicing factor members. **(A)** Diagram of predicted interactions generated in Cyoscape.v3 between miRNAs-*MAPT*-*SRSF* gene family members identified from the miRWalk database. **(B)** Heat map for the Spearman correlation matrix between miRNA expression and mRNA levels of the SRSF factors analyzed. The color of each grid determines the degree of correlation between variables, where positive correlations are shown in red and negative correlations in blue. For each correlation, statistical significance is indicated by the *p-value*: *\* p-value* < 0.05, ** *p-value* < 0.01, *\*\*\* p-value* < 0.001. The exact values of the correlation coefficients used to make the matrix and *p-value* are given in Tables S8 and S9. **(C).** Analysis of the relative mRNA expression levels of several SRSF family members was assayed by RT-qPCR in control subjects (*N =* 4), and LOAD patients (non-*HTT IAs* carriers, *N =* 14; *HTT IAs, N =* 13). GAPDH mRNA was used as normalizer. Relative mRNA expression was performed using the comparative Ct method (2^-ΔΔCt^), considering control group as a reference. The results are shown as the median ± SD. Each point represents one subject. Kruskal-Wallis’s test followed by Dunn’s multiple comparison test were used to calculate statistical significance for *SRSF1/2/3/4/7/9/11*. One-way ANOVA test followed by Tukey’s test was applied for *SRSF6.* **(D)** Representative images of SRSF6 immunoblot. Protein determination was performed in LOAD patients (non-*HTT IAs* carriers, *N =* 7; *HTT IAs, N =* 7). The molecular weight (kDa) is indicated. **(E)** Relative levels of SRSF6, expressed as arbitrary optical density units. Non-*HTT IAs* carriers’ group was used control. Results are shown as mean ± SD. Each point represents one subject. Statistical significance was calculated by Student’s t-test. *P-values* are indicated in the graphs.

Therefore, we wanted to evaluate the expression level of SRSF mRNAs found in caudate samples from the donors included in this study, considering other family members that have also been linked to Tau splicing [22]. The results showed that the expression levels were lower in the vast majority of the analyzed family members of LOAD patients with respect to the control subjects and, in some cases, even more so in LOAD patients carrying *HTT IAs* (Figure 3B). In fact, such downregulation may potentially be exerted by several miRNAs or by only one. For example, *SRSF1* has up to 19 possible interactions from seven of the selected miRNAs, whereas *SRSF4* has only one interaction (miR-9-5p; Figures 3A and 3B). Thus, it will be necessary to determine which miRNAs are the most important in terms of regulation of these SRSFs.

To further explore the relationship between altered miRNAs in the caudate nucleus and *SRSF* expression, we performed a correlation analysis between the 11 differentially expressed miRNAs among LOAD donors and mRNA expression of selected SRSF family members (Figure 3C; Table S8-S9). Negative correlations were observed between the expression of three miRNAs (miR-128-3p, miR-30d-5p, and miR-487-3p) and several *SRSF* mRNAs. Specifically, *SRSF1*, *SRSF2*, *SRSF3*, and *SRSF4* correlated with the three of them; *SRSF11* with miR-128-3p and miR-30d-5p; and *SRSF6* and *SRSF9* only with miR-30d-5p. Therefore, the differential miRNA profile in the caudate nucleus of LOAD patients could be affecting the expression of this family, potentially modifying the expression of isoforms in the *MAPT* gene.

Given that Fernandez-Nogales et al. (2014) had previously demonstrated SRSF6 factor dysfunction in the striatal nucleus of HD patients compared to healthy individuals, and increased levels at both the mRNA and protein levels [10], we investigated SRSF6 levels in our LOAD samples. Although no significant differences in mRNA expression were observed (Figure 3B), we found a significant reduction in SRSF6 protein levels in LOAD patients carrying *HTT IAs* compared to non-carrier (Figures 3C, D; Figure S5). These findings suggested a relationship between the expression of splicing factors and the miRNAs identified. Specifically, for SRSF6, the overexpression of certain miRNAs could be mediating a repression in the translation of its mRNA, resulting in a decrease in protein expression, which contrasts with that previously described in HD.

### FUS and SFPQ protein levels are higher in LOAD patients carrying *HTT IAs*

After studying the expression of SRSF splicing factors in relation to the identified miRNAs, we extended the analysis to other regulatory proteins of the spliceosome. Using *in silico* tools, we identified that five miRNAs, including miR-140-3p, directly target the mRNA of the RNA-binding protein (RBP) FUS (Figure 4A).

**Figure 4.**
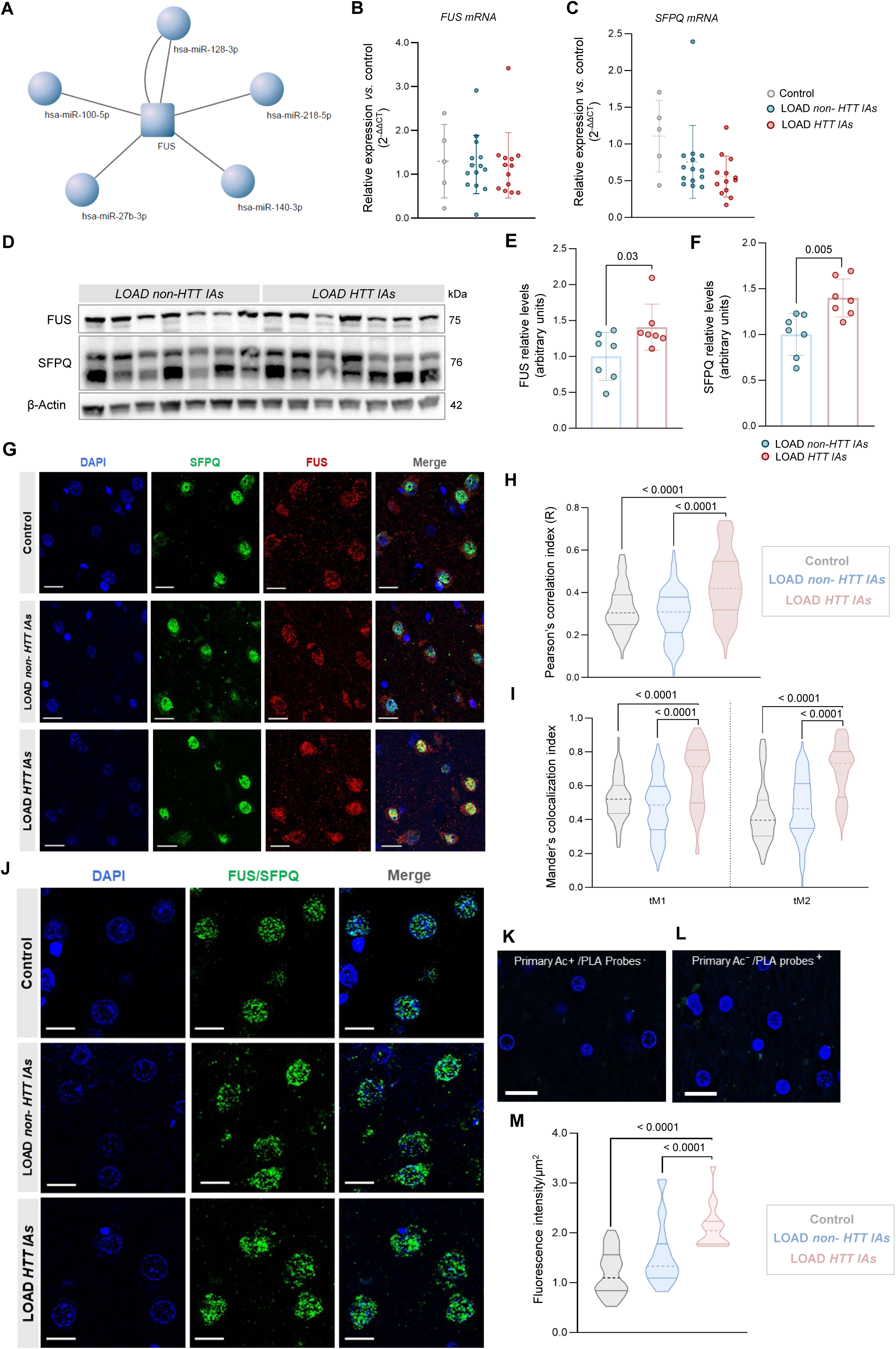
*HTT* Intermediate alleles increases FUS-SFPQ complexes in the caudate nucleus of LOAD patients. **(A)** Cyoscape.v3-generated diagram showing the interactions between miRNAs and their various targets identified from the miRWalk database with the FUS gene. **(B-C)** Relative expression levels of FUS **(B)** and SFPQ **(C)** mRN, analyzed by RT-qPCR, in control subjects (*N =* 4) and LOAD patients (non-*HTT IAs* carriers, *N =* 14; *HTT IAs, N =* 13). GAPDH mRNA was used as normalizer. Calculation of relative mRNA expression was performed using the comparative Ct method (2^-ΔΔCt^), considering control group as a reference. The results are shown as the median ± SD. Each point represents one subject. **(D)** Representative images for FUS and SFPQ immunoblots. Protein analysis was performed in LOAD patients (non-*HTT IAs* carriers, *N =* 7; *HTT IAs, N =* 7). The molecular weight (kDa) is indicated. **(E-F)** Quantification of the relative levels of FUS (**E**) and SFPQ (**F**), expressed as arbitrary optical density units. Non-*HTT IAs* carriers’ group was used control. Results are shown as mean ± SD. Each point represents one subject. For statistical significance, in the case of mRNA expression the Kruskal-Wallis’ test followed by Dunn’s multiple comparison test was used; for protein level by western blot, the Student’s t-test was used. **(G)** Representative double-fluorescence images of signal colocalization between SFPQ and FUS in caudate neuron nuclei from control subjects (*N =* 3) and LOAD (non-*HTT IAs* carriers, *N =* 4; *HTT IAs, N =* 4). Scale bar, 20 µm. **(H)** Representation of Pearson’s correlation coefficient (R) between SFPQ and FUS signal. **(I)** Colocalization analysis using Threshold Mander’s coefficients (tM1 and tM2). The value of tM1 represents the overlap of the SFPQ channel pixels with the FUS channel pixels, and tM2 represents the overlap of the FUS channel pixels with the SFPQ channel pixels. For colocalization analysis, 30 nuclei in each patient randomly selected along six layers of the Z-plane were analyzed by the Coloc2 plugin of the FIJI software. Data are shown as median and interquartile ranges. Each point represents the average value for the six Z-plane layers in a nucleus. Kruskal-Walli’s test followed by Dunn’s multiple comparison test was used to calculate statistical significance. **(J)** Representative images showing PLA signal in nuclei of caudate neurons from control subjects (*N =* 2) and LOAD patients (non-*HTT IAs* carriers, *N =* 2; *HTT IAs, N =* 2). Scale bar, 10 µm. **(K-L)** Negative controls of the PLA technique by omission of hybridization probes and/or primary antibodies. **(M)** Signal intensity quantification by summing the maximum projection of colocalization spots in each nucleus. Data are shown as median and interquartile ranges. Each point corresponds to one nucleus. Statistical significance was calculated by Kruskal-Wallis’ test followed by Dunn’s multiple comparison. *P-values* are indicated in the graphs.

Previous research had identified a nuclear complex between FUS and the splicing factor SFPQ as an essential mechanism for *MAPT* gene splicing, through the formation of an intranuclear dimer that facilitates E10 excision [23]. In certain neurodegenerative disorders, such as tauopathies, this interaction is altered, impacting on the dysregulation of splicing and on the abnormal production of Tau protein isoforms [24]. To elucidate the role of the FUS-SFPQ complex, and its possible modulation by the presence of *HTT IAs*, we assessed the levels of FUS and SFPQ in the caudate nucleus. No differences in FUS and SFPQ mRNA levels were found between the groups studied (Figures 4B, C). However, for both, protein levels were higher in *HTT IA* carrier patients compared to non-carriers (Figures 4D-F; Figure S6). Increased protein levels in FUS and SFPQ could favor the formation, and perhaps also the stability, of the complex, impacting the alternative splicing of E10 in *MAPT* [24]. In this scenario, FUS-SFPQ complex could influence the pathogenesis of LOAD due to their crucial role on Tau splicing.

### Caudate neurons of *HTT IA* carrier patients show increased formation of nuclear FUS-SFPQ complexes

The FUS-SFPQ complex plays a key role in the synthesis of Tau protein isoforms, Tau 4R and Tau 3R, and its disruption causes an imbalance that favors the formation of pure 4R tauopathies, such as FTD [24]. However, in the case of AD, Tau 4R/3R ratio remains the same as in healthy subjects [24]. Therefore, we investigated whether the elevation of FUS and SFPF protein levels observed in LOAD carrier patients involves a change in the formation of FUS-SFPQ complex.

We evaluated the presence of FUS-SFPQ complexes by determining their nuclear distribution and colocalization (Figure 4G). To provide a quantitative measure of the degree of colocalization, we performed two different approaches. First, we used Pearson’s correlation coefficient (R; Figure 4H), which showed similar levels of colocalization between the control group and LOAD patients non-carrying *HTT IAs*, whereas LOAD carrier patients presented significantly higher colocalization. This was further confirmed by determination of Mander’s threshold colocalization coefficients (tM1 and tM2; Figure 4I). To validate these results, we used a specific technique to detect close protein interactions (PLA; Figure 4J-L). Quantification of fluorescence intensity showed a significantly higher signal in the LOAD group with *HTT IAs* compared to controls and LOAD non-carriers, which would correspond with increased complex formation of both proteins (Figure 4M). These findings corroborate previous colocalization results, suggesting that the presence of *HTT IAs* in LOAD patients may increase the interaction between FUS and SFPQ, enhancing the formation of splicing complexes.

### The presence of CAG IAs in the *HTT* gene increases diffuse HTT protein levels in caudate neurons of LOAD patients

It has been previously described that HTT levels are increased in the neurons of hippocampus and frontal cortex patients [12]. To determine whether *HTT IAs* affects HTT protein and its distribution in LOAD patients, a histopathological analysis was performed on caudate nucleus, using samples from HD patients as positive control (Figure 5A). The results showed that the number of HTT-positive neurons in the cytoplasm was higher in both LOAD groups compared with the healthy control group (*p-value* < 0.000, ANOVA; Figure 5B). However, the group carrying *HTT IAs* had an even higher number of positive neurons, compared to non-carriers (*p-value* < 0.0001, ANOVA; Figure 5B). Interestingly, no intranuclear inclusions of HTT (neuropathological hallmark of HD) were found in neurons from LOAD patients, with only diffuse cytoplasmic HTT present in these patients. We performed immunoblot analysis in LOAD donors, which revealed high molecular weight immunoreactive HTT (>250 kDa; Figure S7). This was subsequently confirmed by ubiquitin labeling, revealing a similar pattern. In both cases, protein levels were significantly increased in LOAD donors with *HTT IAs* (Figure S7). All in all, these results were consistent with previously published data [12], extending the regions in which there is an increase of HTT-positive neurons in the brains of LOAD patients. Thus, it is possible that the presence of *HTT IAs* may induce early stages of HTT protein aggregation in the caudate nucleus of LOAD patients. Moreover, it reinforces the role of the *HTT IAs* as potential disease modifiers, as they may act as elements of concurrence of proteinopathies at the brain level.

**Figure 5.**
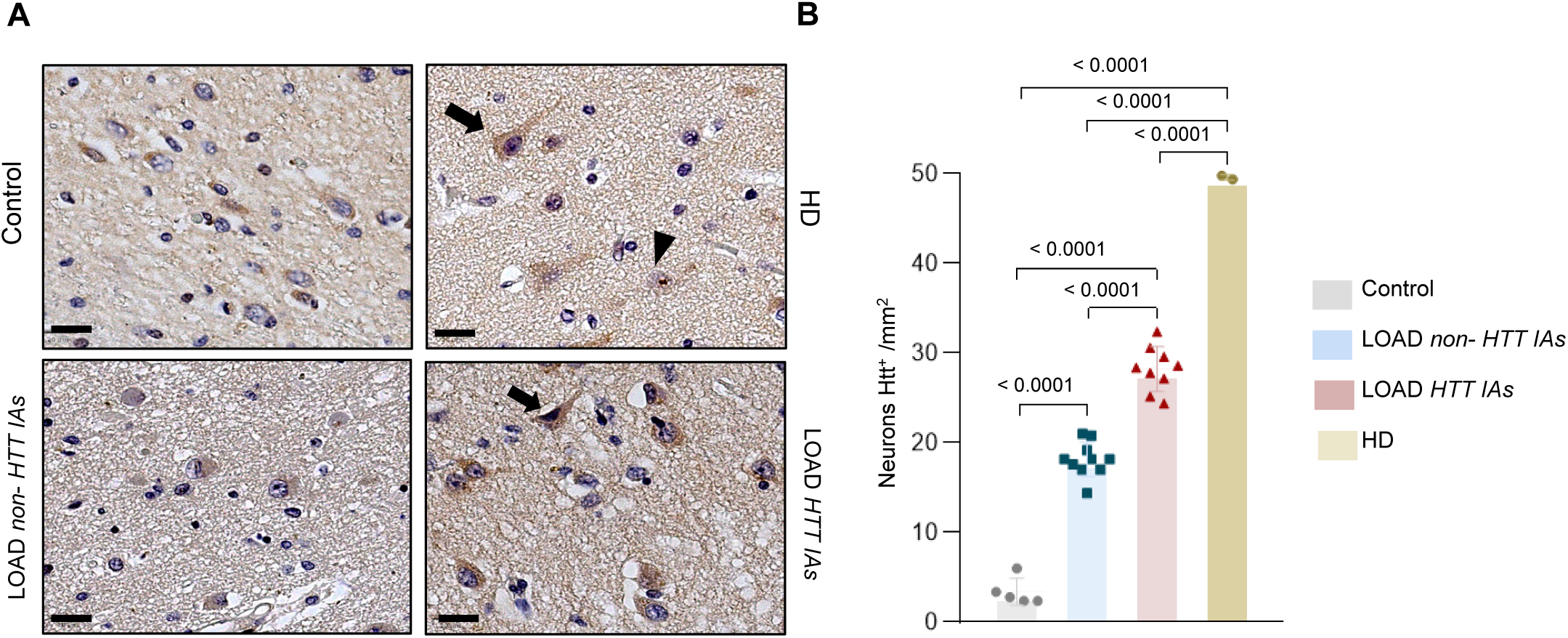
The presence of *HTT IAs* increases the number of caudate nucleus neurons with diffuse HTT. **(A)** Representative images of IHC localization of HTT in caudate neurons of control subjects (*N =* 5), LOAD patients (non-*HTT IAs* carriers, *N =* 10; *HTT IAs, N =* 9), and HD patients (*N =* 2). The arrows in the IAs carrier and HD patient show a cytoplasmic pattern of HTT. The arrowhead in the HD patient shows an intranuclear inclusion of HTT. Scale bar, 20 µm. **(B)** Quantification of HTT protein-positive neurons in the striatal nucleus. Data are shown as mean ± SD. Each point represents one subject. Statistical significance was calculated by one-way ANOVA test followed by Tukey’s test. *P-values* are indicated in the graph.

### Altered splicing due to the presence of *HTT IAs* increases Tau 3R and 4R isoforms in the caudate nucleus of LOAD patients

Based on the found dysfunction in the splicing machinery, mediated by SRSF factors and/or the FUS-SFPQ complex, we investigated mRNA expression levels of *MAPT* and Tau 3R and 4R transcripts in the caudate nucleus of healthy controls and LOAD patients. No significant differences in *MAPT* and Tau 3R levels were observed among the three groups (Figure 6A-B). However, Tau 4R levels were significantly higher in the non-carrier LOAD group compared to controls (Figure 6B). Next, we analyzed the levels of total Tau protein and the 3R and 4R isoforms only in the LOAD groups (Figure 6C; Figure S8). Total Tau levels were significantly higher in carrier patients (Figure 6D), with a trend toward higher levels of the 3R isoform in the *HTT IAs* carrier group (Figure 6E). No changes in Tau 4R levels were detected (Figure 6F). Since, at the protein level, we were detecting increased levels of Tau 3R, it prompted us to analyze how the two Tau isoforms, 3R and 4R, were distributed at the histological level.

**Figure 6.**
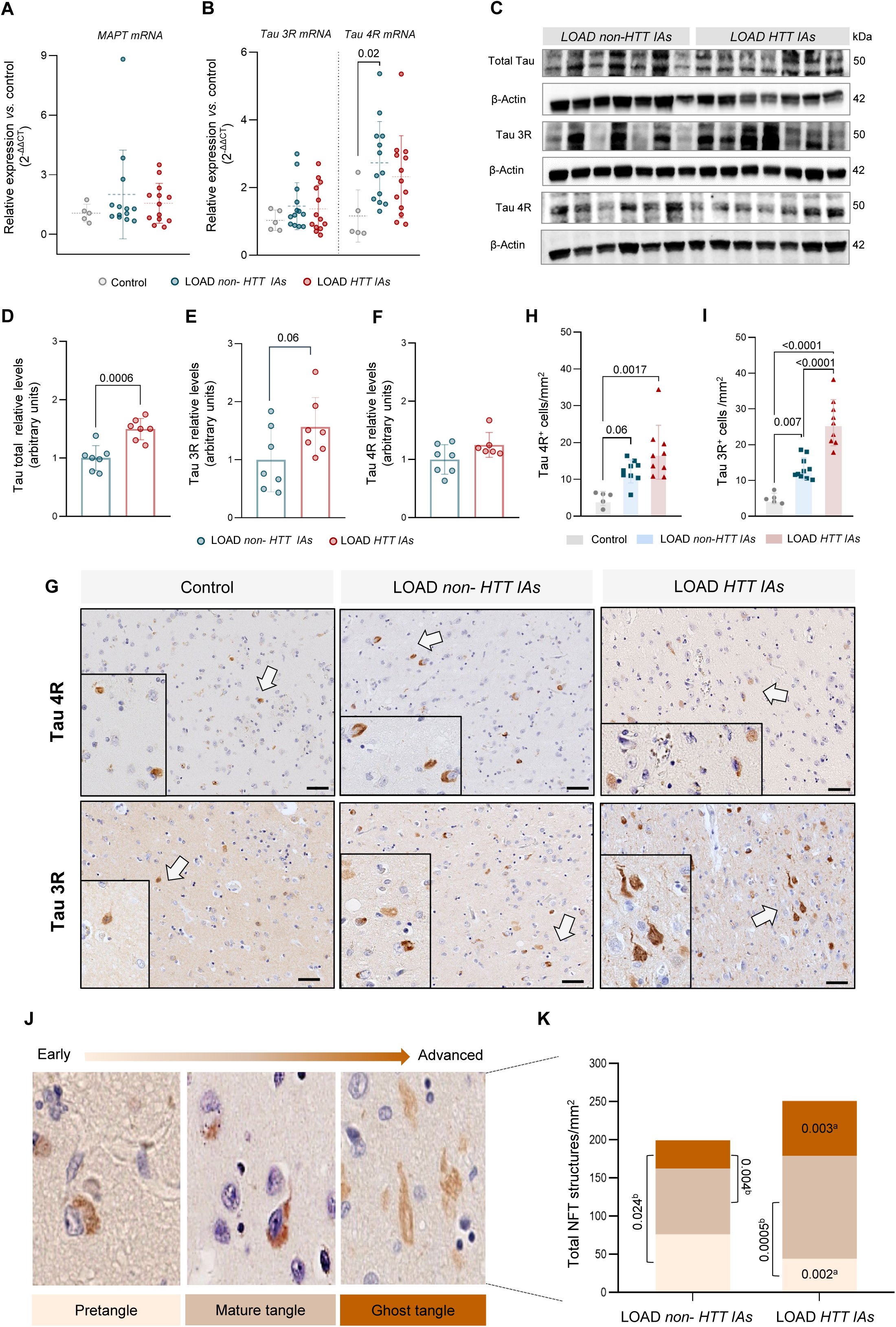
*HTT IAs* increase Tau isoforms and leads to advanced Tau tangles in LOAD patients. **(A-B)** Relative expression levels of *MAPT* gene and Tau 3R and 4R transcripts, respectively, analyzed by RT-qPCR in control subjects (*N =* 4) and LOAD patients (non-*HTT IAs* carriers, *N =* 13; *HTT IAs, N =* 13). GAPDH mRNA was used as normalizer. The comparative Ct method (2 ^-ΔΔCt^) was used for calculation of relative mRNA levels. Data are shown as the median ± SD. statistical significance, in the case of qPCR the Kruskal Wallis’ test followed by Dunn’s multiple comparison test was used **(C)** Representative images for total Tau, Tau 3R, and Tau 4R immunoblots. Protein analysis was performed in LOAD patients (non-*HTT IAs* carriers, *N =* 7; *HTT IAs, N =* 7). The molecular weight (kDa) is indicated. **(D-F)** Quantification of the relative levels of total (**D**), 3R (**E**), and 4R (**F**) Tau, expressed as arbitrary optical density units. **(G)** Representative images of IHC expression of Tau 4R (upper panel) and Tau 3R (lower panel) in striatal neurons from control subject (*N =* 5) and LOAD patients (non-*HTT IAs* carriers, *N =* 9; *HTT IAs, N =* 9). Arrows show the amplified region in each case. Scale bar: 50 µm. **(H-I)** Quantification of positive neurons for Tau 4R (**H**) and Tau 3R (**I**) in the striatal nucleus. Data are represented as mean ± SD. Each point represents one subject. **(J)** Representative images of total NFT structures quantified in IHC images for Tau 4R and Tau 3R in LOAD donors (non-*HTT IAs* carriers, *N =* 9; *HTT IAs, N =* 9). **(K)** Quantification of NFT structures in neurons of the caudate nucleus. Data are shown as the total structures for Tau 4R and Tau 3R in each case. Statistical analysis: Kruskal-Wallis’ test followed by Dunn’s multiple comparison test (A-B), Student’s t-test (D-F, K^a^), one-way ANOVA test followed by Tukey’s *post hoc* test (H-I, K^b^). *P-values* are indicated in the graph.

Distribution and density of Tau 4R and 3R isoform-positive neurons in different regions of the caudate nucleus in healthy subjects and in both LOAD groups is shown in Figure 6G. Quantification of the 4R isoform revealed a higher number of Tau 4R^+^ neurons in LOAD patients compared to controls, although only significant in the case of *HTT IAs* carriers (Figure 6H). Regarding Tau 3R isoform, both LOAD groups presented a higher number of Tau 3R^+^ neurons compared to controls, with a more pronounced increase in LOAD subjects carrying *HTT IAs* (Figure 6I). These results suggest that, in the caudate nucleus, the presence of *HTT IAs* affects tau pathology by altering the synthesis of the 4R and 3R isoforms, possibly due to incorrect alternative *MAPT* mRNA processing that disturbs the balance between the isoforms.

### Advanced Tau tangles predominate in LOAD patients with *HTT IAs* in a CAG expansion size-dependent manner

Neurofibrillary tangles (NFTs), key elements in the pathophysiology of AD, begin as fibrillar bundles in neurons, evolve into mature tangles, and are externalized after neuronal death [25, 26]. This evolution is unidirectional and correlates with the differential manifestation of Tau 4R/3R isoforms. Ghost tangles, the final stage (Figure 6J), show predominantly the 3R isoform, indicating a chronological shift from 4R to 3R during the progression of AD [27]. Therefore, to examine whether there is a shift from 4R to 3R profile, we quantify the number of NFTs structures in both groups of LOAD patients.

We first analyzed the proportion of each stage of NFT maturation within the same group of LOAD patients. Non-carrier subjects exhibited a higher number of pretangle-positive cells and mature tangles compared to ghost tangles (Figure 6K). Conversely, patients with HTT IAs showed a reduced pretangle count alongside an increased presence of mature tangles, with no significant differences observed in ghost tangles (Figure 6K).

When comparing NFTs structures between the two groups, the primary differences emerged at the early and late stages of maturation. Specifically, LOAD patients with HTT IAs displayed fewer pretangles and a greater number of ghost tangles than the non-carrier group (Figure 6K). These findings suggest that the presence of *HTT IAs* is associated with a faster neuropathological progression, impacting Tau aggregation. This may indicate a more accelerated neurodegenerative process in LOAD patients carrying *HTT IAs*.

Finally, in order to represent a translational view of the results to the clinical environment, we analyzed the relationship between miRNA expression, CAG expansion size, as well as HTT-positive neurons, and the status of NFTs aggregates (Figure 7; Table S10). We observed a positive association between all miRNAs, except miR-9-3p and miR-100-5p, and the number of HTT-positive neurons. The percentage of pretangles shows a statistically significant negative correlation with 7 of the 11 miRNAs, whereas only three of them were positively correlated with ghost tangles. Furthermore, CAG expansion size reflected a statistically significant correlation with the number of HTT-positive neurons and the advanced state of Tau aggregates, ghost tangles, while the correlation was inverse with the number of pretangles. These results highlight the key role of *HTT IAs* as epigenetic regulators, with an effect that adds to the presence of the disease itself on miRNAs. This not only has a relevant impact on the development of Tau protein pathology, but also emphasizes the role of HTT in facilitating more accelerated neurodegeneration.

**Figure 7.**
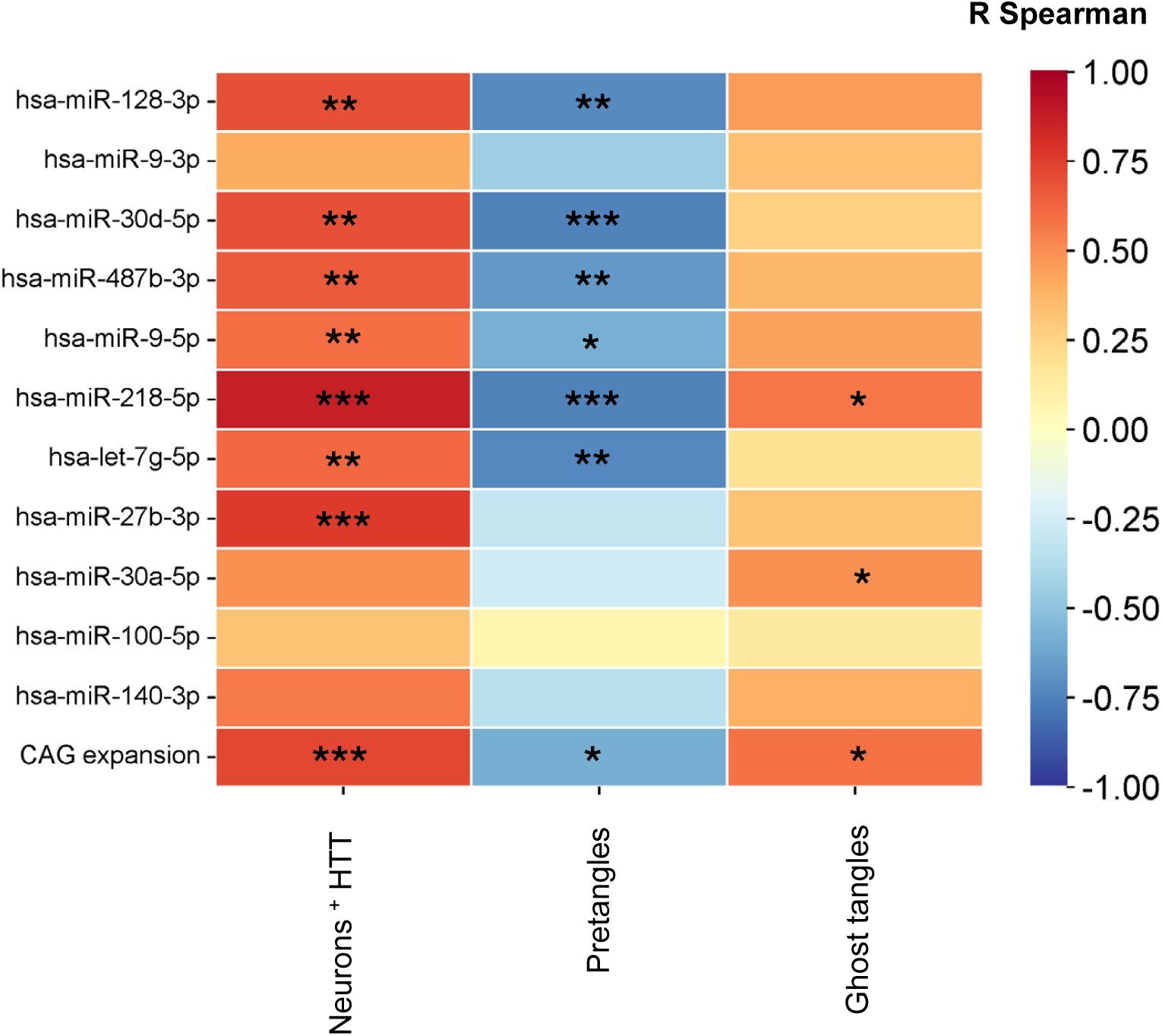
*HTT IAs* as epigenetic regulators. Heat map for the Spearman correlation matrix between miRNA levels and CAG expansion size *vs.* histopathological markers related to Tau and HTT. The color of each grid determines the degree of correlation between variables, where positive correlations are shown in red and negative correlations in blue. For each correlation, statistical significance is indicated by the *p-value*: *\* p-value* < 0.05, ** *p-value* < 0.01, *\*\*\* p-value* < 0.001. The exact values of the correlation coefficients used to make the matrix and *p-value* are given in Table S10.

## Discussion

Evidence suggests that proteinopathies may have a common pathophysiological pathway, since it is no longer surprising to find, at the neuropathological level, aggregates of other altered proteins, apart from the pathogenic protein causing the primary neurological disease. Among the proteins most frequently described in copathologies, the Tau protein, alpha-synuclein and TDP-43 [1, 2] stand out. There are even studies that suggest as risk factors for this phenomenon not only age, but also the presence of the APOE4 isoform [1], highly associated with the development of LOAD [28].

Along with all this, there are several studies where the possible pathogenic effect of CAG repeats, characteristic of polyglutaminopathies, have been evaluated in different ND. For example, the size of the CAG repeat in the *ATXN1* gene is associated with different clinical features in AD, such as memory, attention and medial temporal lobe atrophy [44]. Likewise, our previous studies have described a higher frequency of CAG repeats within the intermediate range in the *HTT* gene in patients with neuropathological diagnosis of certain alpha-synucleinopathies and tauopathies, most notably AD [4, 5]. Mazzeo et al. observed in patients with subjective cognitive impairment that the presence of *HTT IA*s interacts with age and APOE4 polymorphism, increasing the probability of progressing to mild cognitive impairment, the preclinical stage of AD [45]. However, the role that these alleles might have on the progression of LOAD has not been addressed so far. Therefore, in this study we aimed to focus on the analysis of the influence of the presence of *HTT IA*s in a group of subjects with a neuropathological diagnosis of LOAD. Surprisingly, at the clinical level, patients carrying *HTT IA*s have lower survival after disease onset, something that had not been previously described. Therefore, *HTT IA*s not only affect stages prior to AD [29], but also could be favoring LOAD to progress more rapidly, so they could be considered as key risk factors/modifiers of the disease.

To determine how *HTT IA*s could accelerate LOAD progression, we focused on the analysis of the miRNAs profile in the caudate nucleus, as this is the region most sensitive to changes in the number of CAG repeats in the *HTT* gene [14]. This profile is altered in LOAD patients with respect to healthy subjects, something that is characteristic of this pathology [30], although it was not described in the caudate nucleus before. We found that all altered miRNAs were overexpressed in LOAD patients compared to healthy subjects and, surprisingly, this overexpression was even higher in LOAD patients carrying *HTT IA*s. Interestingly, only one miRNA was different between the two groups of patients, miR-140-3p. Thus, this genetic condition affects epigenetic regulatory mechanisms more powerfully than in the case of non-carrier LOAD patients. This finding underscores the relevance of exploring the role of miRNAs in LOAD, as it could shed light on how the presence of these *HTT IA*s further influences altered molecular pathways in LOAD patients. Indeed, using *in silico* tools, we found that the 11 miRNAs overexpressed in LOAD patients carrying *HTT IA*s in the caudate nucleus regulate key pathways in neurodegeneration and affect common pathways in NDs. All this reinforces the hypothesis that there are shared pathogenic events in proteinopathies [2].

One of the most relevant pathways that we have been able to identify is that of the spliceosome, as it is directly involved in Tau pathology [46]. In fact, the most altered miRNAs target several members of the splicing factor family SRSF, and their epigenetic regulation has been previously described (reviewed in [47]). Thus, we determined that several splicing factors were down-regulated and that this was related to an increase in the expression of miRNAs, such as miR-128-3p, miR-30d-5p and miR-487b-3p. Considering that changes in the profile of miRNAs in the caudate nucleus were more exacerbated in patients carrying *HTT IA*s, it makes sense that in cases such as SRFS1, SRSF4, SRSF9 and, to a lesser extent, SRSF3, expression levels were lower in this group of patients. However, for the rest of the SRSFs that we have analyzed and that did not show changes, regulation could occur at the protein level, possibly by translation blockage [48]. Such was the case of the SRSF6 factor, known for its interaction with the *HTT* gene in HD subjects, since it showed lower protein levels in LOAD donors carrying *HTT IA*s. This splicing factor is involved in *MAPT* gene processing, promoting the 4R isoform of Tau [10, 49], as are the SRSF1 and SRSF9. This could justify the trend we have observed in terms of Tau 3R levels. Therefore, the presence of *HTT IA*s could contribute to the increase in Tau 3R, promoting an alteration in the 4R/3R balance, traditionally described in AD neuropathology. To further support this shift towards Tau 3R observed in LOAD patients carrying *HTT IA*s, we found an increased presence of FUS/SFPQ complexes in these subjects. This could further favor the displacement of *MAPT* E10 splicing towards a predominant production of Tau 3R. Therefore, this fact not only depends on the action of SRSF factors but could also involve this splicing regulatory complex at the nuclear level.

Tau 3R isoform, unlike Tau 4R, could destabilize the axonal cytoskeleton and reduce neuronal lifespan [31], leading to a possible cellular vulnerability in LOAD patients with *HTT IAs*. In fact, the increased presence of the Tau 3R in the caudate nucleus stands out as a relevant finding, since previous literature indicates that high levels of Tau 3R favor the accelerated progression of NFTs towards their last stage, ghost tangles, promoting further neuronal damage [32]. In our study, LOAD patients with *HTT IAs* showed a lower percentage of pretangle and a significant increase in ghost tangles compared to non-carrier patients, which could explain the decrease in survival observed in these patients after diagnosis. In parallel, we were able to determine the existence of a higher number of diffuse cytoplasmic HTT-positive neurons in the caudate nucleus in LOAD patients (as previously described for the hippocampus and cortex, [12]). Moreover, these levels are significantly higher in the subset of LOAD patients carrying *HTT IAs*. These findings suggest that, at least in this brain region, HTT could acquire aberrant conformations that affect neuronal physiology, potentially contributing to accelerated disease progression through altered *MAPT E10* splicing.

In summary, LOAD patients show an altered miRNA profile in the caudate nucleus, not previously described, and the presence of *HTT IAs* further exacerbates the differences with respect to healthy subjects. This profile affects key components of the spliceosome, such as SRSF family factors and the nuclear FUS-SFPQ complex, which accelerates both Tau 3R production and NFT maturation. Moreover, the presence of *HTT IAs* not only promotes faster maturation of NFTs, but also increases the presence of soluble HTT protein in neurons. This dual impact suggests a copathological state that requires future research to further explore the functional consequences of this interaction and to identify possible therapeutic strategies aimed at mitigating these alterations.

## Conclusions and limitations

Our findings, supported by previous studies, confirm a link between intermediate CAG expansions in the *HTT* gene and LOAD. In particular, we highlight for the first time the profile of miRNAs in LOAD caudate nucleus as essential regulators in the molecular mechanisms underlying disease progression. The accelerated progression in Tau pathology together with the presence of soluble HTT protein reinforces the existence of a copathology condition, where the interaction between different pathological proteins may accelerate neurodegeneration. Taken together, the epigenetic alterations and their histopathological consequences identified in this study could open new perspectives for the development of targeted therapeutic strategies, which could be particularly relevant for this subgroup of LOAD patients. These findings do not only contribute to increase our understanding of the physiopathology of the disease and its clinical heterogeneity, but can also have important and direct implications in terms of future personalized medicine. As *HTT* genotyping is a widely used technique, our results may contribute to a better segmentation and homogenization of groups of patients in clinical trials for AD and better and future clinical practice. However, we are aware that our study has several limitations, since the sample size of the patient groups in some analyses was small, mainly due to the difficulty in the proposed methodologies, so it is necessary to replicate this study with a larger set of subjects in some respects. Furthermore, given that the patients included in our study are classified as Braak stage V-IV, a very advanced stage of the disease, another relevant aspect would be to evaluate whether patients with early (I-II) or moderate (III-IV) Braak stage show the same alteration in the miRNAs profile and, consequently, in *MAPT* alternative splicing. This will allow us to understand in depth whether the dysfunction occurs at the beginning or changes progressively during the progression of the disease. It would also be necessary to study classical regions of AD progression (hippocampus or cortex areas) to verify whether the effect of *HTT IAs* is the same or, as they seem to be more resistant to HTT changes [14], whether resilience in areas other than the caudate is greater.

## Abbreviations

AD: Alzheimer’s disease

*APOE*: Apolipoprotein E

E10: exon 10 of *MAPT* gene

FTD: frontotemporal dementia

FUS: fused in sarcoma protein

HD: Huntington’s disease

*HTT IAS*: intermediate alleles in *HTT* gene

HTT: huntingtin protein

KEEG: Kyoto Encyclopedia of Genes and Genomes

LOAD: Late-onset Alzheimer’s disease

*MAPT*: microtubule associated Tau gene

mHTT: mutant huntingtin

miRNAs: microRNAs

NDs: neurodegenerative diseases

NFTs: neurofibrillary tangles

PCA: principal component analysis

PLA: proximity ligation assay

PMI: *postmortem* interval

polyQ: enlarged polyglutamine tract

RBP: RNA binding protein

RIN: RNA integrity number

RPM: read per million

RT: room temperature

SFPQ: Proline/Glutamine rich splicing factor

SRSF: Serine/Arginine rich splicing factor

## Data Availability

All data produced in the present study are available upon reasonable request to the authors. The databases obtained and used during the current study are available on ZENODO, with the accession number “11185109” (https://doi.org/10.5281/zenodo.11185109).

## Acknowledgments

We are indebted to the patients and their families. Authors wish to thank Asociación *Parkinson Asturias-Obra Social Cajastur* for their support. We would like to sincerely thank the Scientific and Technical Services at the University of Oviedo, especially the Microscopy and Image Processing Unit, for their valuable support. We also appreciate the Molecular Histopathology Service in Animal Cancer Models at Instituto Universitario de Oncología del Principado de Asturias (IUOPA) and the Molecular Genetics Laboratory of Hospital Universitario Central de Asturias. We want to particularly acknowledge for its collaboration, the Principado de Asturias BioBank (PT23/0077), financed jointly by Servicio de Salud del Principado de Asturias, Instituto de Salud Carlos III with European funds Also, we are indebted to the HCB-IDIBAPS Biobank for sample and data procurement. Finally, we would like to thank Javier Bermejo-Pampliega for his work as a research technician at the University of Oviedo, supported by a contract associated with grant AYUD/2021/5134 from FICYT, confounded by the Fondo Europeo de Desarrollo Regional (FEDER).

## Author’s contribution

J.C-S performed all histological and molecular experiments, conducted data analysis, created the figures and wrote the manuscript. S.P-G performed and analyzed the clinical data and genotyping the initial samples to establish the studied cohort. P.P-H, M.F-S and E.I-G provided guidance on the analysis and interpretation of RNAseq and miRNAs experiments. MD.C-T developed the technical tasks for the preparation of the samples provided by the Biobank of the Principality of Asturias. V.A and M.M-G conceptualized, designed and obtained funding for the study, interpreted the epidemiological data, executed the genetic objective of the study, and supervised the final versions of the manuscript. C.T-Z designed and supervised the molecular and histopathological analysis experiments, interpreted the data, and supervised the different versions of the manuscript.

## Declarations

### Ethical approval

All procedures were performed after obtained the approval of Ethical Committees of HCB-IDIBAPS Biobank (A1-C23001; Barcelona, Spain), and Ethics Committee of the Principality of Asturias (CEImPA n° 2022.266).

### Consent for publication

All authors have approved the content of this manuscript and provided consent for publication.

