## Supplementary material for "Synergistic impact of CAG intermediate alleles in the *HTT* gene and microRNA dysregulation exacerbates spliceosome impairment and accelerates Tau pathology in the caudate nucleus of late-onset Alzheimer’s disease"

Table S1. Clinical and neuropathological information for selected subjects to RNAseq and histological analysis

| Patient ID | Biobank | Gender | Onset age | Death age | Braak stage | APOE | Disease duration | HTT short allele | HTT long allele | P.M.I (h) |
|---|---|---|---|---|---|---|---|---|---|---|
| **Control** | | | | | | | | | | |
| CT1.1 | BPA | M | *N. A* | 80-85 | *N. A* | 3/3 | *N. A* | 18 | 21 | 5:30 |
| CT1.2 | BPA | F | *N. A* | 70-75 | *N. A* | 3/3 | *N. A* | 18 | 21 | 4:50 |
| CT1.3 | BPA | M | *N.A* | *76-80* | *N.A* | 3/4 | *N.A* | 17 | 17 | *N.A* |
| CT1.4 | BPA | M | *N. A* | 66-70 | *N. A* | 2/4 | *N. A* | 15 | 23 | 4:10 |
| CT1.5 | BPA | M | *N. A* | 66-70 | *N. A* | 3/3 | *N. A* | 17 | 26 | 4:00 |
| CT1.6 | BPA | F | *N. A* | 70-75 | *N. A* | 3/3 | *N. A* | 15 | 17 | 7:10 |
| **LOAD *non-HTT IAs*** | | | | | | | | | | |
| AD1.1 | BPA | M | *N. A* | 66-70 | VI | 4/4 | *N. A* | 18 | 20 | 4:00 |
| AD2.1 | IBIBAPS | M | 70-75 | 86-90 | VI | 3/3 | 14 | 15 | 18 | 10:00 |
| AD2.2 | IDIBAPS | M | 76-80 | 86-80 | V | 3/4 | 9 | 16 | 18 | 15:00 |
| AD2.3 | IDIBAPS | F | 70-75 | 76-80 | VI | 3/4 | 6 | 17 | 18 | 10:00 |
| AD2.4 | IDIBAPS | M | 76-80 | 80-85 | VI | 3/3 | 6 | 17 | 17 | 8:45 |
| AD2.5 | IDIBAPS | F | 76-80 | 80-85 | V | 3/4 | 4 | 11 | 19 | 6:00 |
| AD2.6 | ISIBAPS | F | 81-85 | 86-90 | V | 3/3 | 7 | 17 | 17 | 13:00 |
| AD2.7 | IDIBAPS | M | 60-65 | 70-75 | V | 4/4 | 9 | 18 | 19 | 6:45 |
| AD2.8 | IDIBAPS | F | 70-75 | 80-85 | V | 3/4 | 14 | 16 | 17 | 5:00 |
| AD2.9 | IDIBAPS | F | 76-80 | 86-90 | VI | 3/4 | 7 | 16 | 17 | 4:30 |
| AD2.10 | IDIBAPS | M | 60-65 | 76-80 | VI | 2/3 | 15 | 17 | 19 | 7:00 |
| AD2.11 | IDIBAPS | F | *N. A* | 80-85 | VI | 2/4 | *N. A* | 15 | 17 | 16:30 |
| AD2.12 | IDIBAPS | M | *N. A* | 70-75 | VI | 4/4 | *N. A* | 17 | 17 | 8:00 |
| AD2.13 | IDIBAPS | F | 76-80 | 86-90 | V | 3/3 | 9 | 16 | 17 | 19:30 |
| **LOAD *HTT IAs*** | | | | | | | | | | |
| AD1.2 | BPA | M | *N. A* | 86-90 | III | 3/3 | *N. A* | 19 | 27 | *N. A* |
| AD1.3 | BPA | M | *N. A* | 60-65 | VI | *N. A* | *N. A* | 17 | 29 | 5:00 |
| AD1.4 | BPA | F | *N. A* | 86-90 | V | 3/3 | *N. A* | 21 | 29 | 4:55 |
| AD2.14 | IDIBAPS | F | *N. A* | 86-90 | VI | 3/3 | *N. A* | 17 | 30 | 13:00 |
| AD2.15 | IDIBAPS | F | 76-80 | 80-85 | V | 3/4 | 3 | 17 | 34 | 5:30 |
| AD2.16 | IDIBAPS | M | 66-70 | 70-75 | V | 3/3 | 5 | 15 | 27 | 6:30 |
| AD2.17 | IDIBAPS | F | 86-90 | 90-95 | V | 3/3 | 7 | 15 | 39 | 12:30 |
| AD2.18 | IDIBAPS | M | 66-70 | 70-75 | VI | 3/3 | 4 | 20 | 30 | 14:30 |
| AD2.19 | IDIBAPS | F | 70-75 | 80-85 | VI | 3/3 | 9 | 18 | 27 | 5:00 |
| AD2.20 | IDIBAPS | M | 60-65 | 76-80 | VI | 2/3 | 15 | 17 | 29 | 5:00 |
| AD2.21 | IDIBAPS | F | *N. A* | 80-85 | VI | 3/3 | *N. A* | 17 | 27 | 5:00 |
| AD2.22 | IDIBAPS | F | 66-70 | 70-75 | VI | 4/4 | 6 | 17 | 29 | 14:00 |
| AD2.23 | IDIBAPS | M | 80-85 | 86-90 | V | 3/4 | 6 | 13 | 27 | 3:00 |

Abbreviations: *N. A*, not available; M, Male; F, Female; PMI, *post mortem* interval; BPA, Biobanco del Principado de Asturias; IDIBAPS, Instituto de Investigaciones Biomédicas August Pi i Sunyer.

Table S2. RNA related aspects to analysed samples.

| | **Control (*N* = 5)** | **LOAD non-*HTT* *IAs* (*N* = 14)** | **LOAD *HTT* *IAs* (*N* = 13)** | ***p-value*** |
|---|---|---|---|---|
| **RNA ng/µl** | | | | |
| Mean (± SD) | 319.61 (136.58) | 231.01 (52.2) | 322.87 (98.18) | *n. s* |
| Range | 148.8-463.5 | 141.7-293.4 | 211.6-583.3 | |
| **RNA ratio 260/280** | | | | |
| Mean (± SD) | 1.95 (0.08) | 1.97 (0.02) | 1.97 (0.04) | *n. s* |
| Range | 1.73-2.02 | 1.92-1.99 | 1.94-2.01 | |
| **RIN value** | | | | |
| Mean (± SD) | 3.65 (0.72) | 3.2 (1.26) | 3.73 (0.61) | *n. s* |
| Range | 2.4-4.9 | 1.5-4.9 | 2.1-4.3 | |
| **28S/18S ratio** | | | | |
| Mean (± SD) | 2.55 (1.47) | 6.76 (1.45) | 2.9 (1.54) | < 0.001[a] <0.001[b] |
| Range | 0.8-5.3 | 4.7-8.7 | 1.3-5.7 | |
| **PMI (h)** | | | | |
| Mean (± SD) | 7:49 (4.27) | 5:08 (1.28) | 9:34 (4.75) | *n. s* |
| Range | 3:00-14:30 | 4:00-7:10 | 4:00-19:30 | |

Abbreviations: n.s, no significance; RIN, RNA integrity number; PMI, *post mortem* interval.
Statistical analysis: One-way ANOVA followed by Tukey's test.
[a]LOAD *HTT* IAs *vs.* Control.
[b]LOAD non-*HTT* IAs *vs.* Control.

| Table S3. Genes validated for qPCR (Anygene) | |
|---|---|
| **Genes** | **NM Code** |
| *SRSF1* | NM_006924.5 |
| *SRSF2* | NM_001195427.2 |
| *SRSF3* | NM_003017.5 |
| *SRSF4* | NM_005626.5 |
| *SRSF7* | NM_005626.5 |
| *SRSF9* | NM_003769.3 |
| *SRSF11* | NM_001350605.2 |
| *SFPQ* | NM_005066.3 |
| *FUS* | NM_004960.3 |
| *β-ACTIN* | NM_001101.5 |
| *GAPDH* | NM_002046.7 |
| *HTT*: huntingtin gene; *SRSF*: serine/arginine *splicing* factor gene; *SFPQ*: proline/glutamine *splicing* factor gene; *FUS*: fused protein in sarcoma gene; *β-ACTIN*: Beta Actin gene; GAPDH: glyceraldehyde-3-phosphate dehydrogenase gene. | |

| Tabla S4. Pre-designed sequences used for qPCR | | |
|---|---|---|
| **Genes** | **Sequence** | **Amplification protocol** |
| *Tau 4R* (forward) | 5′ GGTGCAGATAATTAATAAGAAGCTGGA 3’ | 10 min 95 °C<br>(40x) 10 s 95 °C<br>30 s 60 °C<br>1 min 60 °C<br>30 s 65 °C |
| *Tau 4R* (reverse) | 5′ GTGTTTGATATTATCCTTTGAGCCAC 3′ | |
| *Tau 3R* (forward) | 5′ GAAGAATGTCAAGTCCAAGATCGG 3′ | |
| *Tau 3R* (reverse) | 5′ GACTATTTGCACCTTCCCGC 3′ | |
| *MAPT* (forward) | 5′AGAGTCCAGTCGAAGATTGGGTC 3′ | |
| *MAPT* (reverse) | 5′ GGGTTTCAATCTTTTTATTTCCTCC 3′ | |

Table S5. Antibodies used for western blot analysis

| Antibody (clone) | Source | Dilution | Vender | Reference |
|---|---|---|---|---|
| Anti- Tau RD3 (8E6/C11) | Mouse | 1:500 | Merck Millipore | 05-803 |
| Anti-Tau RD4 (1E1/A6) | Mouse | 1:500 | Merck Millipore | 05-804 |
| Anti-FUS/TLS (4H11) | Mouse | 1:500 | Santa Cruz Biotechnology | sc-47711 |
| Anti- SFPQ (B92) | Mouse | 1:1000 | Abcam | Ab11825 |
| Anti-Tau total (TAU-5) | Mouse | 1:400 | Invitrogen | AHB0042 |
| Anti-SRSF6 | Rabbit | 1:500 | LSBio | LS-B5712 |
| Anti- HTT (EM-48) | Mouse | 1:500 | Merck Millipore | MAB5374 |
| Anti-Ubiquitina | Rabbit | 1:500 | Abcam | Ab7780 |

Table S6. Frequency of APOE isoforms in cohorts studied.

| APOE alleles | Control (*N* = 335) | LOAD (*N* = 323) | *p-value*[a] |
|---|---|---|---|
| Ɛ2, Ɛ2 | 1 (0.30%) | 1 (0.31%) | 0.746 |
| Ɛ2, Ɛ3 | 40 (12.01%) | 4 (1.23%) | <0.001 |
| Ɛ2, Ɛ4 | 9 (2.70%) | 10 (3.09%) | 0.484 |
| Ɛ3, Ɛ3 | 221 (66.36%) | 151 (46.74%) | <0.001 |
| Ɛ3, Ɛ4 | 60 (18.01%) | 127 (39.31%) | <0.001 |
| Ɛ4, Ɛ4 | 2 (0.60%) | 29 (8.97%) | <0.001 |

Abbreviations: LOAD, late onset Alzheimer's disease.

[a]Data are shown as N (%). Statistical analysis: Fisher's test.

| Table S7. Loading coefficient of each miRNA in the different PCs. | | | |
|---|---|---|---|
| PC1 contribution | | PC2 contribution | |
| miRNA | Loading | miRNA | Loading |
| **miR-9-5p** | 0.880789 | **miR-125b-5p** | 0.606046 |
| **miR-30a-5p** | 0.849922 | **miR-221-3p** | 0.453611 |
| **miR-30d-5p** | 0.779295 | **miR-30c-5p** | 0.437675 |
| **miR-218-5p** | 0.767932 | **miR-103a-3p** | 0.392599 |
| **miR-126-3p** | 0.76425 | **miR-29a-3p** | 0.351416 |
| **miR-16-5p** | 0.757481 | **miR-128-3p** | 0.342787 |
| **miR-103a-3p** | 0.735448 | miR-143-3p | 0.330433 |
| **miR-29a-3p** | 0.735443 | **miR-30d-5p** | 0.318899 |
| **let-7g-5p** | 0.728606 | **miR-126-3p** | 0.308592 |
| miR-26a-5p | 0.726799 | **miR-487b-3p** | 0.285647 |
| **miR-27b-3p** | 0.721046 | miR-335-5p | 0.272675 |
| **miR-9-3p** | 0.713063 | **miR-30a-5p** | 0.254821 |
| **miR-221-3p** | 0.713043 | **miR-24-3p** | 0.246767 |
| **miR-487b-3p** | 0.710713 | **miR-30e-5p** | 0.117875 |
| **miR-30e-5p** | 0.698314 | **miR-99a-5p** | 0.107254 |
| **miR-24-3p** | 0.692466 | **miR-100-5p** | -0.02751 |
| **miR-128-3p** | 0.690724 | **miR-9-5p** | -0.0552 |
| miR-140-3p | 0.680506 | **let-7a-5p** | -0.09241 |
| let-7f-5p | 0.680058 | miR-21-5p | -0.10347 |
| **miR-99a-5p** | 0.676334 | **miR-9-3p** | -0.11184 |
| **miR-100-5p** | 0.674124 | miR-7-5p | -0.13343 |
| miR-101-3p | 0.666458 | **miR-16-5p** | -0.13798 |
| miR-26b-5p | 0.658125 | **miR-181a-5p** | -0.24132 |
| **miR-30c-5p** | 0.654343 | **miR-218-5p** | -0.29567 |
| **let-7a-5p** | 0.650969 | **miR-27b-3p** | -0.31174 |
| **miR-181a-5p** | 0.63889 | **let-7g-5p** | -0.31676 |
| miR-21-5p | 0.619503 | **miR-124-3p** | -0.3663 |
| miR-335-5p | 0.594056 | miR-140-3p | -0.37447 |
| **miR-125b-5p** | 0.579251 | miR-101-3p | -0.38433 |
| **miR-124-3p** | 0.520711 | let-7f-5p | -0.47165 |
| miR-143-3p | 0.295522 | miR-26b-5p | -0.56633 |
| let-7i-5p | 0.248941 | miR-26a-5p | -0.57746 |
| miR-7-5p | 0.156292 | let-7i-5p | -0.74693 |

| Table S8. Spearman correlation coefficient between miRNA expression, CAG expansion number and histopathological hallmarks analyzed. | | | | | | | | |
|---|---|---|---|---|---|---|---|---|
| Rho Spearman | *SRSF1* | *SRSF2* | *SRSF3* | *SRSF4* | *SRSF6* | *SRSF7* | *SRSF9* | *SRSF11* |
| miR-128-3p | -0.65 | -0.62 | -0.56 | -0.46 | -0.38 | -0.40 | -0.40 | -0.46 |
| miR-9-3p | 0.05 | -0.16 | -0.06 | -0.07 | 0.29 | 0.16 | 0.10 | 0.18 |
| miR-30d-5p | -0.65 | -0.59 | -0.67 | -0.65 | -0.46 | -0.44 | -0.50 | -0.47 |
| miR-487b-3p | -0.61 | -0.57 | -0.57 | -0.47 | -0.40 | -0.38 | -0.38 | -0.31 |
| miR-9-5p | -0.05 | -0.18 | -0.14 | -0.14 | -0.12 | 0.01 | 0.02 | -0.04 |
| miR-218-5p | -0.33 | -0.28 | -0.23 | -0.18 | -0.35 | -0.11 | -0.11 | -0.09 |
| let-7g-5p | -0.27 | -0.34 | -0.27 | -0.25 | -0.38 | -0.17 | -0.13 | -0.40 |
| miR-27b-3p | -0.04 | 0.08 | 0.07 | 0.11 | 0.09 | 0.29 | 0.25 | 0.20 |
| miR-30a-5p | -0.11 | -0.06 | -0.15 | -0.18 | 0.02 | -0.07 | -0.11 | 0.07 |
| miR-100-5p | 0.03 | 0.11 | 0.23 | 0.15 | 0.42 | 0.44 | 0.35 | 0.09 |
| miR-140-3p | -0.01 | 0.00 | 0.17 | 0.15 | 0.22 | 0.40 | 0.35 | 0.02 |

| Table S9. Related *p-value* corresponding to the spearman correlation between miRNA expression, the number of CAG expansions and the histopathological characteristics analyzed. | | | | | | | | |
|---|---|---|---|---|---|---|---|---|
| *p-value* | *SRSF1* | *SRSF2* | *SRSF3* | *SRSF4* | *SRSF6* | *SRSF7* | *SRSF9* | *SRSF11* |
| miR-128-3p | 0.0003 | 0.0005 | 0.0025 | 0.0152 | 0.0516 | 0.0384 | 0.0411 | 0.0166 |
| miR-9-3p | 0.8206 | 0.4148 | 0.7715 | 0.7120 | 0.1380 | 0.4345 | 0.6365 | 0.3589 |
| miR-30d-5p | 0.0003 | 0.0012 | 0.0001 | 0.0003 | 0.0152 | 0.0226 | 0.0085 | 0.0126 |
| miR-487b-3p | 0.0007 | 0.0018 | 0.0019 | 0.0129 | 0.0384 | 0.0536 | 0.0507 | 0.1102 |
| miR-9-5p | 0.8088 | 0.3720 | 0.4906 | 0.5003 | 0.5400 | 0.9711 | 0.9110 | 0.8277 |
| miR-218-5p | 0.0899 | 0.1522 | 0.2586 | 0.3556 | 0.0742 | 0.5853 | 0.5686 | 0.6387 |
| let-7g-5p | 0.1796 | 0.0809 | 0.1755 | 0.2126 | 0.0516 | 0.3991 | 0.5120 | 0.0404 |
| miR-27b-3p | 0.8300 | 0.6850 | 0.7393 | 0.5811 | 0.6628 | 0.1467 | 0.2068 | 0.3091 |
| miR-30a-5p | 0.5769 | 0.7484 | 0.4509 | 0.3573 | 0.9350 | 0.7370 | 0.5874 | 0.7279 |
| miR-100-5p | 0.8703 | 0.5980 | 0.2520 | 0.4602 | 0.0274 | 0.0226 | 0.0747 | 0.6496 |
| miR-140-3p | 0.9783 | 0.9879 | 0.3854 | 0.4546 | 0.2680 | 0.0375 | 0.0710 | 0.9086 |

Table S10. Spearman correlation coefficient and *p-value* between miRNA expression, CAG expansion number and histopathological hallmarks analyzed.

| | **HTT$^+$ Neurons** | | **Pretangles** | | **Ghost tangles** | |
|---|---|---|---|---|---|---|
| | Rho Spearman | *p-value* | Rho Spearman | *p-value* | Rho Spearman | *p-value* |
| miR-128-3p | 0.69 | 0.0011 | -0.72 | 0.0008 | 0.45 | 0.0615 |
| miR-9-3p | 0.40 | 0.0905 | -0.45 | 0.0594 | 0.33 | 0.1762 |
| miR-30d-5p | 0.69 | 0.0011 | -0.75 | 0.0004 | 0.26 | 0.2892 |
| miR-487b-3p | 0.66 | 0.0021 | -0.67 | 0.0024 | 0.36 | 0.1468 |
| miR-9-5p | 0.60 | 0.0065 | -0.58 | 0.0112 | 0.43 | 0.0745 |
| miR-218-5p | 0.86 | 0.0000 | -0.75 | 0.0003 | 0.57 | 0.0131 |
| let-7g-5p | 0.61 | 0.0060 | -0.73 | 0.0006 | 0.18 | 0.4731 |
| miR-27b-3p | 0.76 | 0.0001 | -0.31 | 0.2060 | 0.32 | 0.2000 |
| miR-30a-5p | 0.49 | 0.0319 | -0.26 | 0.2909 | 0.49 | 0.0402 |
| miR-100-5p | 0.32 | 0.1767 | 0.07 | 0.7724 | 0.14 | 0.5812 |
| miR-140-3p | 0.56 | 0.0125 | -0.35 | 0.1555 | 0.39 | 0.1145 |
| CAG expansion | 0.72 | 0.0006 | -0.59 | 0.0101 | 0.58 | 0.0119 |

*Supplementary figures for:*

### Synergistic impact of intermediate alleles in the *HTT* gene and microRNA dysregulation exacerbates spliceosome impairment and accelerates Tau pathology in the caudate nucleus of late-onset Alzheimer's disease

Juan Castilla-Silgado[1,2,6], Sergio Perez-Oliveira[2,4,6], Paola Pinto-Hernandez[1,2], Manuel Fernandez-Sanjurjo[1,2], Maria Daniela Corte-Torres[2], Eduardo Iglesias-Gutierrez[1,2], Manuel Menendez-Gonzalez[2,3,5], Victoria Alvarez[2,4,#,*], Cristina Tomas-Zapico[1,2,#].

[1] Department of Functional Biology (Physiology), University of Oviedo, 33006, Oviedo, Spain.

[2] Instituto de Investigación Sanitaria del Principado de Asturias (ISPA), 33011, Oviedo, Spain.

[3] Department of Neurology, Hospital Universitario Central de Asturias (HUCA), 33011, Oviedo, Spain.

[4] Genetics laboratory, Hospital Universitario central de Asturias, 33011, Oviedo, Spain.

[5] Department of Medicine, University of Oviedo, 33006, Oviedo, Spain.

[6] Parkinson association of Asturias, 33011, Oviedo, Spain.

[7] Biobank of Principado de Asturias, Hospital Universitario Central de Asturias (HUCA), 33011, Oviedo, Spain.

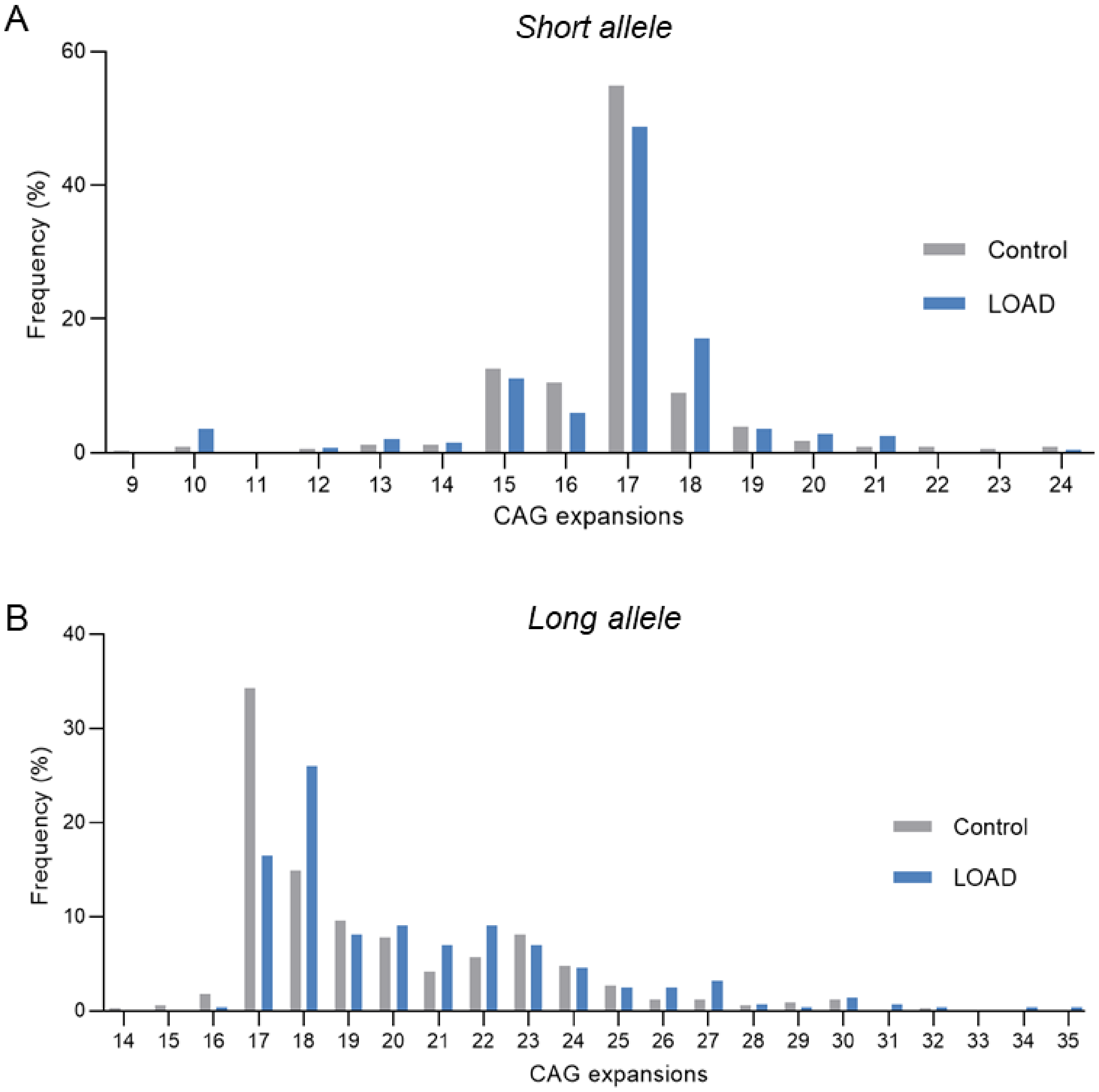


**Figure S1. Frequency of CAG expansions in the *HTT* gene in control and LOAD groups.** **A**. Within the minor allele of the *HTT* gene, a high frequency of 17 repeats was observed in both groups, with no expansion found in the intermediate range. **B**. The distribution of CAG expansions in the major allele revealed that the most frequent number was 17 in the control group and 18 in LOAD patients. Both groups showed a homogeneous distribution within the nonpathological range of 17-24 CAGs, although there are cases of expansions above 27 CAGs. Abbreviations: LOAD, late-onset Alzheimer's disease; *HTT*, huntingtin gene.

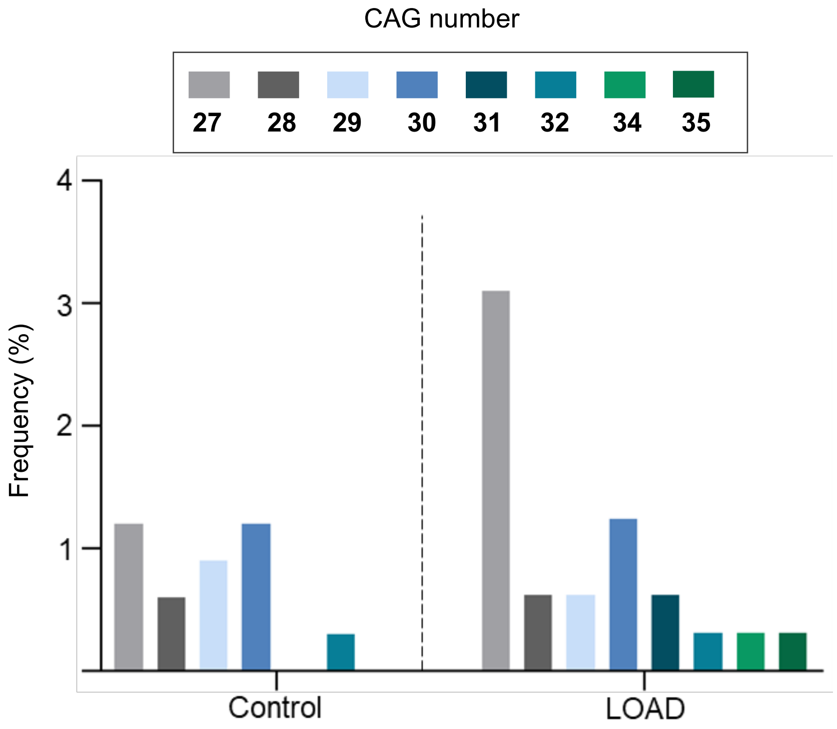


**Figure S2. Intermediate CAG expansions distribution in the *HTT* gene in control subjects and LOAD patients**. Control subjects presented a homogeneous distribution with a very low frequency, with 30 being the highest number of repeats observed. In contrast, LOAD patients showed repetitions throughout the intermediate range (27-35), with 27 being the most frequent number. Abbreviations: LOAD, late-onset Alzheimer's disease.

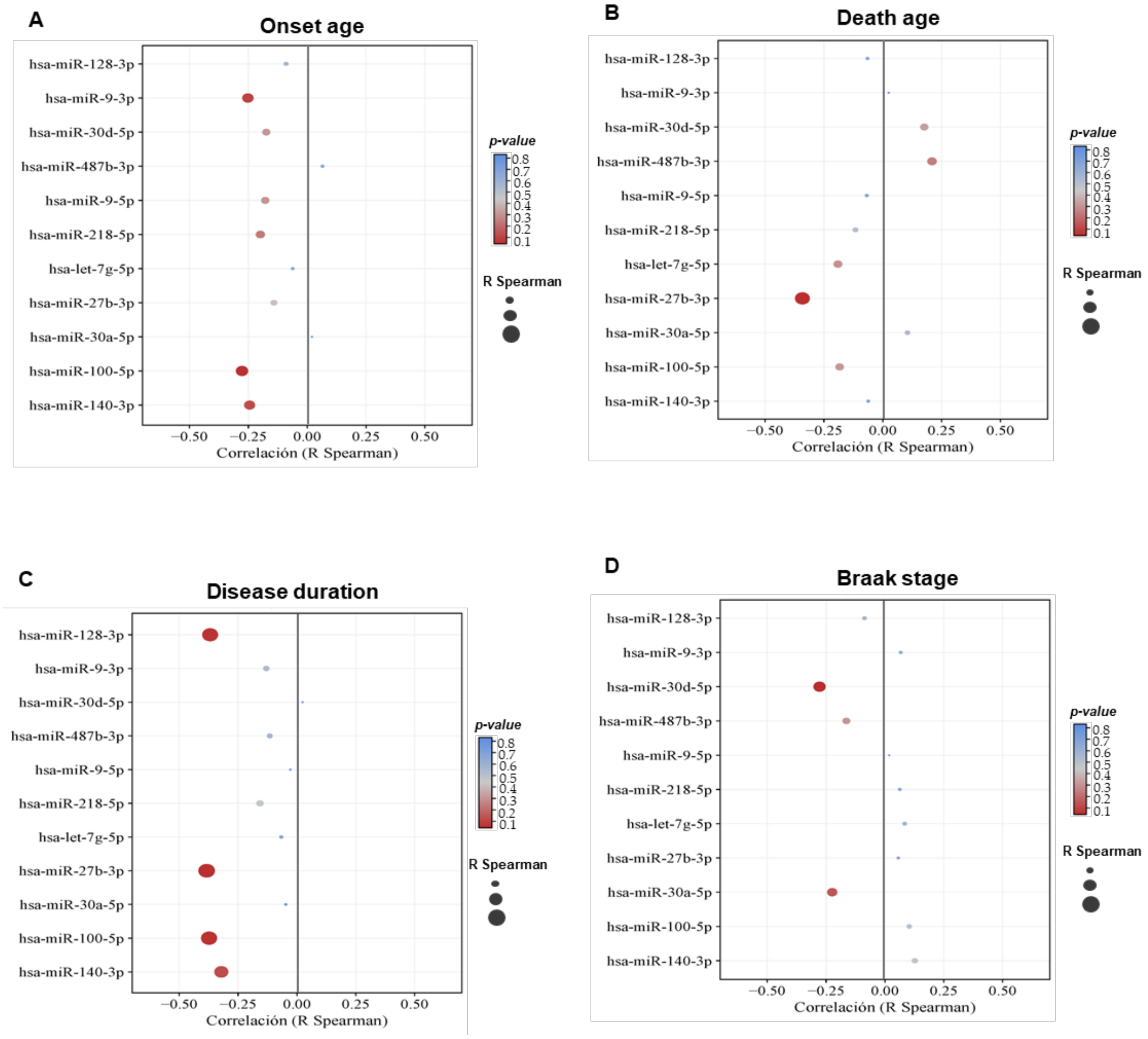


**Figure S3. Spearman's Rho correlation between recorded clinical variables and miRNA array expression. A**. Age at disease onset. **B**. Recorded age at death. **C.** Total duration of disease in years. **D.** Braak stage. The size of the circle in each correlation is proportional to the Spearman's R value and the color scale represents the level of significance given by the *p-value*.

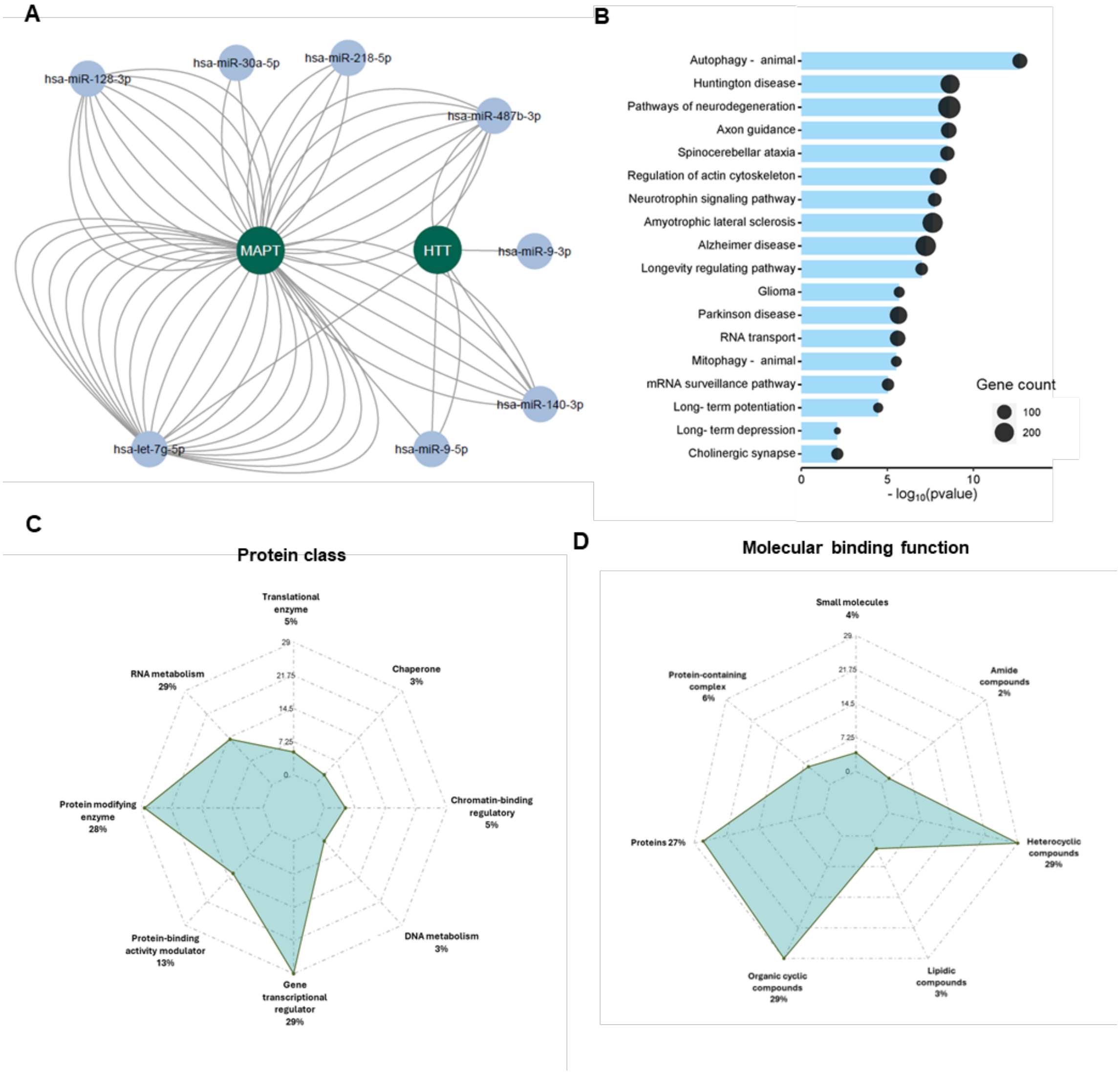


**Figure S4. In silico analysis of interactions and predicted pathways for altered miRNAs in the caudate nucleus. A.** Diagram of interactions generated in Cyoscape.v3 between significant miRNAs and their various targets identified from the miRWalk database between MAPT and HTT genes**. B.** Diagram of enriched pathways targeting differentially expressed miRNAs between LOAD groups *vs*. controls. **C-D**. Distribution of the percentage of target genes classified in Panther DB v18.0 according to protein type and predominant molecular function.

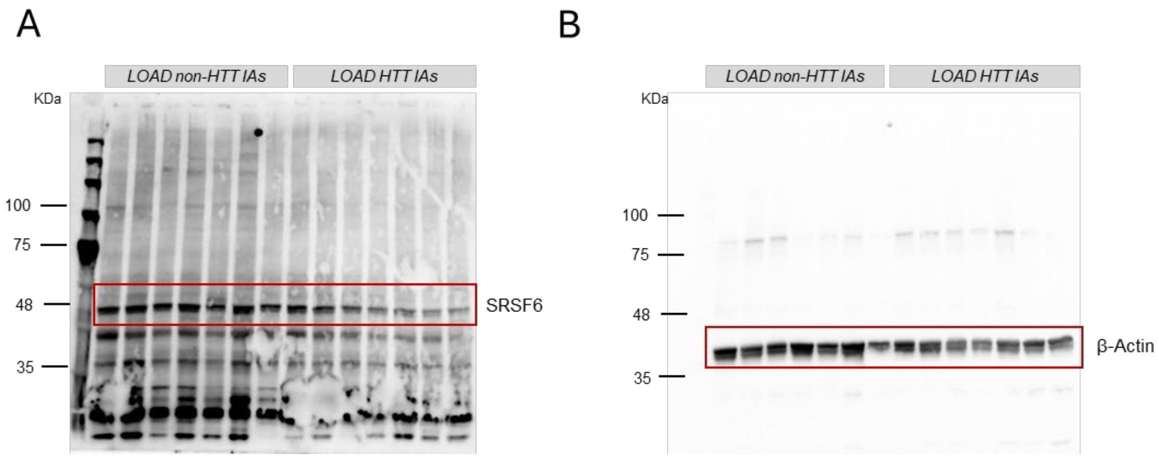


**Figure S5. Uncropped western blot image in figure 3. A**. SRSF6 protein expression**. B.** β-actin protein expression. Red boxes correspond to the cropped area to the main figure.

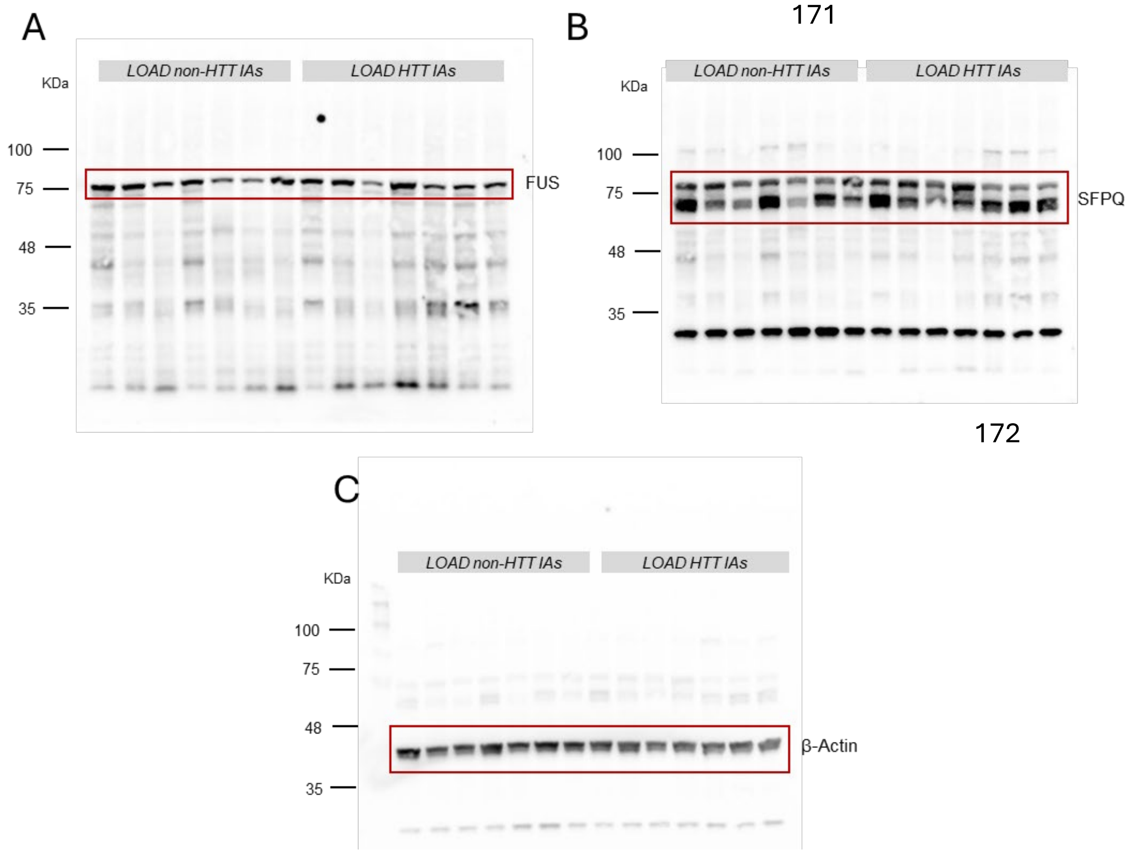


**Figure S6. Uncropped western blot image in figure 4. A**. FUS protein expression**. B.** SFPQ protein expression. **C.** β-actin protein expression. Red boxes correspond to the cropped area to the main figure.

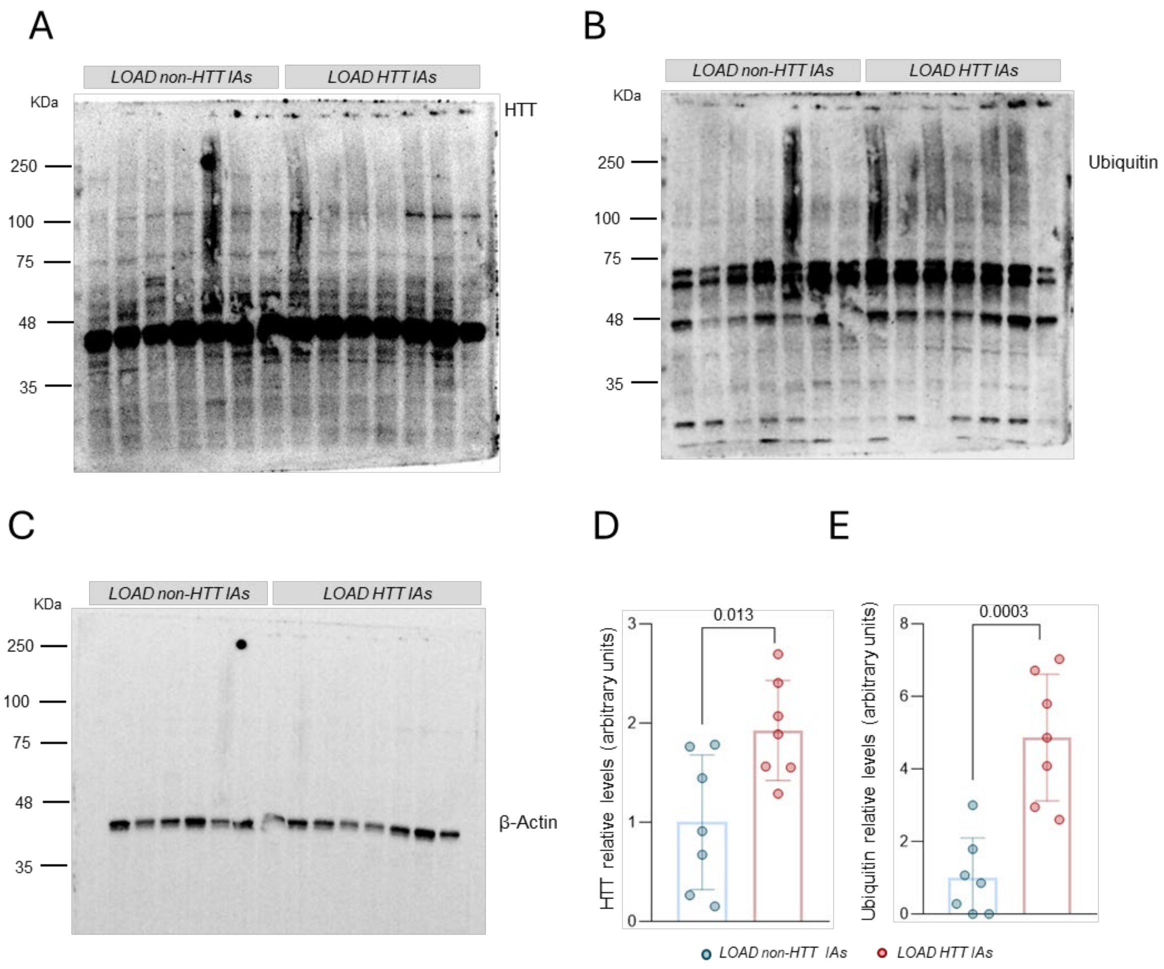


**Figure S7. HTT and ubiquitin protein level in caudate nucleus of LOAD patients. A**. HTT protein expression**. B.** Ubiquitin protein expression. **C.** β-actin protein expression. **D.** Quantification of immunoreactive structures with anti-HTT antibody. **E.** Quantification of immunoreactive structures with anti-ubiquitin antibody.

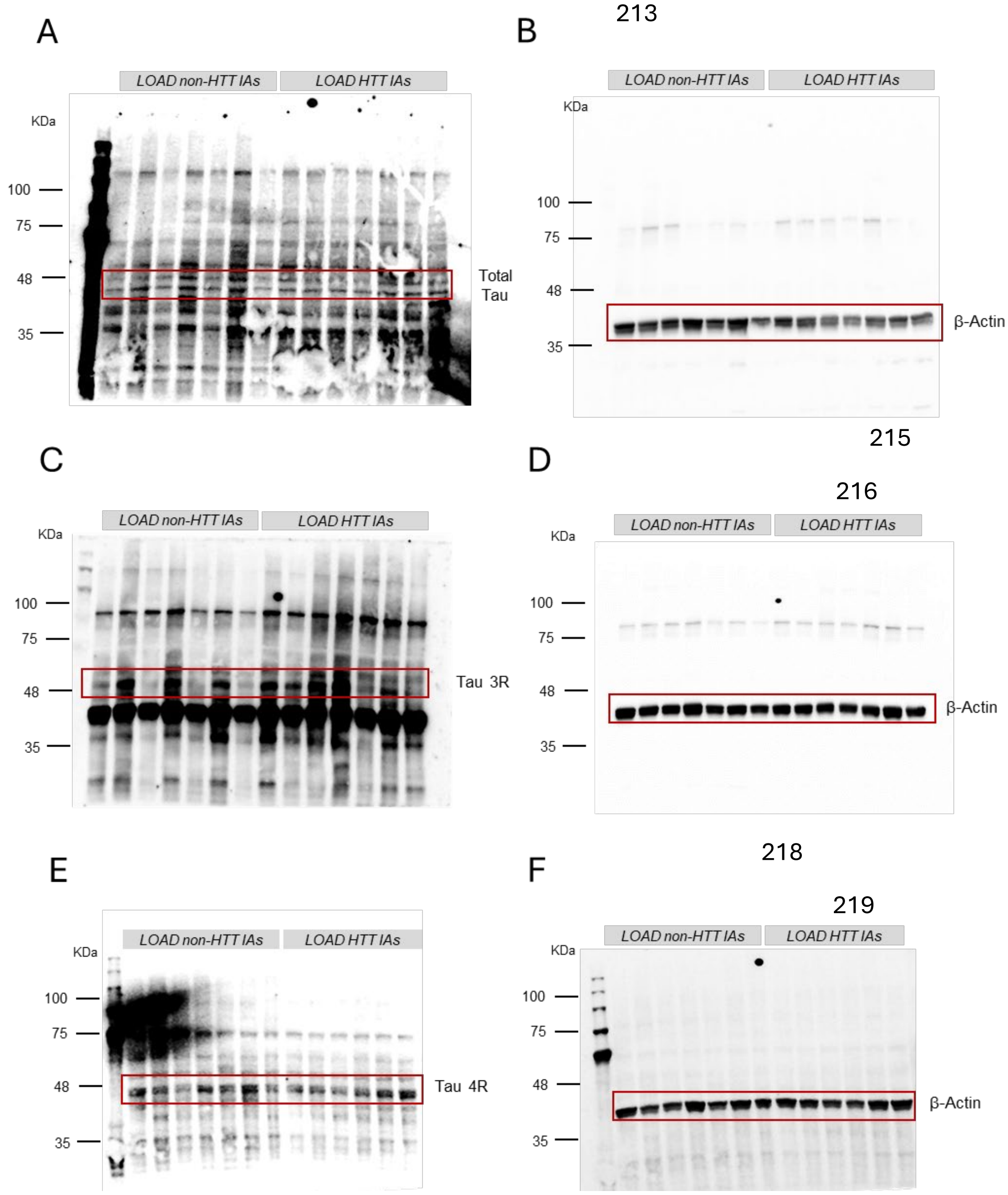


**Figure S8. Uncropped western blot image in figure 6. A**. Total Tau protein expression**. B.** β-actin protein expression in Total Tau membrane **C.** Tau 3R protein expression. **D.** β-actin protein expression in Tau 3R membrane. **E.** Tau 4R protein expression. **F.** β-actin protein expression in Tau 4R membrane. Red boxes correspond to the cropped area to the main figure.
